# Comparison of the stability of Glycoprotein Acetyls and high sensitivity C-reactive protein as markers of chronic inflammation

**DOI:** 10.1101/2023.03.02.23286349

**Authors:** Daisy C.P. Crick, Golam M Khandaker, Sarah L Halligan, David Burgner, Toby Mansell, Abigail Fraser

**Affiliations:** Population Health Sciences, Bristol Medical School, University of Bristol, Bristol, UK; MRC Integrative Epidemiology Unit at the University of Bristol, Bristol, UK; Department of Psychology, University of Bath, Bath, UK; Department of Psychiatry and Mental Health, University of Cape Town, South Africa; Department of Psychiatry, Stellenbosch University, South Africa; Murdoch Children’s Research Institute, Royal Children’s Hospital, Parkville, Victoria, Australia; Department of Paediatrics, Melbourne University, Parkville, Victoria, Australia; NIHR Bristol Biomedical Research Centre, Bristol, UK; Avon and Wiltshire Mental Health Partnership NHS Trust, Bristol, UK; Centre for Academic Mental Health, University of Bristol, Bristol, UK

**Keywords:** Inflammation, Glycoprotein Acetyls, C-reactive Protein, Biomarker, Stability, ALSPAC

## Abstract

**Background:** It has been suggested that glycoprotein acetyls (GlycA) better reflects chronic inflammation than high sensitivity C-reactive protein (hsCRP), but paediatric/life-course data are sparse.

**Method:** Using data from the Avon Longitudinal Study of Parents and Children and UK Biobank, we compared short-(over weeks) and long-term (over years) correlations of GlycA and hsCRP, cross-sectional correlations between GlycA and hsCRP, and associations of pro-inflammatory risk factors with GlycA and hsCRP across the life-course.

**Results:** GlycA showed high short-term (weeks) stability at 15y (r=0.75; 95% CI=0.56, 0.94), 18y (r=0.74; 0.64, 0.85), 24y (r=0.74; 0.51, 0.98) and 48y (r=0.82 0.76, 0.86) and this was comparable to the short-term stability of hsCRP at 24y. GlycA stability was moderate over the long-term, for example between 15y and 18y r=0.52; 0.47, 0.56 and between 15y and 24y r=0.37; 0.31, 0.44. These were larger than equivalent correlations of hsCRP. GlycA and concurrently measured hsCRP were moderately correlated at all ages, for example at 15y (r=0.44; 0.40, 0.48) and at 18y (r=0.55; 0.51, 0.59).

We found similar associations of known proinflammatory factors and inflammatory diseases with GlycA and hsCRP. For example, BMI was positively associated with GlycA (mean difference in GlycA per standard deviation change in BMI=0.08; 95% CI=0.07, 0.10) and hsCRP (0.10; 0.08, 0.11).

**Conclusion:** This study showed that GlycA has greater long-term stability than hsCRP, however associations of proinflammatory factors with GlycA and hsCRP were broadly similar.

**Key messages:**

- GlycA is a novel composite biomarker of inflammation which may have greater stability compared to commonly used biomarkers of inflammation such as hsCRP.
- GlycA has comparable short-term stability, but greater long-term stability compared to hsCRP.
- The associations between proinflammatory factors and CRP and GlycA are similar.

## Introduction

Acute inflammation is triggered by microbial infection and factors such as noxious stimuli and tissue injury. It is characterised by the rapid increase of inflammatory-related markers (e.g., acute-phase proteins and cytokines) locally and systemically, and by the accumulation of immune cells in affected tissue ^1 2^. A dysregulation of the immune system can prevent the resolution of the acute inflammatory response and lead to a state of systemic, low-grade chronic inflammation ^3–5^. Certain factors, which can be biological, social or behavioural can also lead to chronic inflammation ^3–5^. Inflammation increases with age ^6^ and differs between the sexes, possibly reflecting differential susceptibility to inflammation-related diseases ^7–9^. Additionally, chronic inflammation is associated with lower socioeconomic position (SEP) ^10^, obesity ^11–13^ and a high alcohol consumption ^14^.

Chronic inflammation is usually assessed using circulating biomarkers such as cytokines and acute-phase proteins and it is associated with the prevalence, incidence and progression of non-communicable diseases ^15^ such as cardiovascular disease (CVD) ^16, 17^ and other non-communicable diseases ^18–26^. For example, a systematic review and meta-analysis of 29 prospective population-based studies found that a one standard deviation increase in the cytokines interleukin-6 (IL-6), interlukin-18 (IL-18) and tumour necrosis factor alpha (TNF-α) was associated with a 10-25% increase in the risk of non-fatal and fatal myocardial infarction ^27^.

A key challenge for inflammation research is identifying biomarkers that reliably reflect systemic, chronic, and low-grade inflammation. Currently, the most widely used biomarker is high sensitivity C-reactive protein (hsCRP), an acute-phase protein ^28^. Other commonly used biomarkers include cytokines such as IL-6 and TNF-α ^29^. However, short-term variability in circulating levels and rapid kinetics in response to infection or other acute stimuli undermine the utility of these single-protein biomarkers as measures of chronic inflammation ^30–32^. Composite biomarkers derived from multiple proteins may be more suitable than single-protein biomarkers when measuring chronic inflammation; unlike single-protein markers, if one component changes, the others may be unaffected, meaning the signal is less subject to acute fluctuations ^31^.

Glycoprotein acetyls (GlycA) is a composite biomarker of inflammation ^33, 34^. It is quantified using Nuclear Magnetic Resonance (NMR) ^35^ and reflects the extent and complexity of N-glycosylation of a number acute-phase proteins ^36^. In response to acute inflammation, acute-phase proteins increase in concentration and glycan complexity, which in turn increases the amplitude of the GlycA NMR signal. However, given that GlycA is a composite marker of inflammation, it should be less reactive to acute environmental changes and, as such, be a more stable measure of chronic inflammation compared to acute-phase markers such as hsCRP ^37^.

GlycA has been shown to be positively associated with inflammation-related non-communicable diseases (NCDs) ^38–40^ such as incident CVD events ^41^. Despite this, studies comparing the short- and long-term stability of GlycA and hsCRP across the life-course are lacking, as are data on associations of GlycA with different pro-inflammatory factors across the life-course.

Here we use data from two large UK population-based cohorts: the Avon Longitudinal Study of Parents and Children (ALSPAC) and the UK Biobank (UKB) to investigate: 1) short-term (weeks) and long-term (years) stability of GlycA levels; 2) correlations between concurrently measured GlycA and hsCRP in adolescence, early-adulthood and mid-adulthood; 3) associations of inflammation-related factors (e.g., smoking) with GlycA and hsCRP and 4) associations of autoimmune/inflammatory diseases (e.g., Crohn Disease) with GlycA and hsCRP. We hypothesised that GlycA would show greater stability over time than hsCRP and display similar or stronger associations with inflammation-related factors (particularly chronic inflammatory states) compared to hsCRP, given that GlycA should reflect chronic inflammation, whereas hsCRP responds to acute inflammatory triggers.

## Materials and Methods

### ALSPAC

The ALSPAC recruited a total of 14,541 pregnant women residing in Avon, UK, with an expected delivery date between 1st April 1991 and 31st December 1992 ^42–44^. The initial number of pregnancies enrolled was 14,541 with 13,988 children alive at 1 year of age. The sample was bolstered and the total sample size for analyses using any data collected after the age of seven was 15,447 pregnancies (14,901 were alive at 1 year of age). Further information about ALSPAC and its ethical guidelines are presented in the supplement.

Offspring were included in this study if they had one measure of GlycA and one measure of hsCRP across ages 15y, 18y and 24y (n=5356; supplementary Figure 1). Mothers were included in this study if they had measures of GlycA and hsCRP at mean age 47y or 50y (n=4480; supplementary Figure 2). After exclusion, no no-white participants remained in the study.

A subset of the participants (N=78-124) who had biological measures repeated for quality control (QC) purposes within 5-6 weeks were used to investigate short-term stability. QC measures for GlycA were available at all clinics but were only available at the 24y clinic for hsCRP.

#### Biomarkers of Inflammation

GlycA and hsCRP were measured in plasma at mean ages 15y, 18y and 24y in the ALSPAC offspring and at mean ages 47y and 50y in the ALSPAC mothers. Details of blood collection, processing and storage, together with biomarker assays are shown in the supplement.

In the maternal cohort, we created combined variables using measures from either the 47y clinic or the 48y clinic (see supplement) separately for the GlycA and hsCRP measures to increase sample size and statistical power.

#### Potential determinants of inflammation

In analyses using offspring data we used SEP, age, BMI, atopy score and sex, symptoms of infection, smoking frequency and alcohol intake as potential determinants of inflammation. In analyses using the maternal cohort we used SEP, age, BMI and atopy score as potential determinants of inflammation. A description of how these variables were recorded/coded is reported in the supplement.

### UK Biobank

The UKB is a community-based, prospective study (https://www.ukbiobank.ac.uk). Recruitment of ∼500,000 participants and baseline assessments were completed between 2006-2010. Participants were included in this study if they had a measure of GlycA and hsCRP (n=112424). A further description of UKB is presented in the supplement.

#### Biomarkers of inflammation

GlycA and hsCRP were measured at the baseline clinic (mean age = 48y) and plasma GlycA was quantified using the same method as in ALSPAC. See supplement for details of sample processing. As in the ALSPAC analysis, hsCRP was converted from mg/L to mmol/L.

#### Potential determinants of inflammation

SEP was indexed using self-reported highest education qualification, participants’ sex, age, BMI, smoking frequency and alcohol intake were included. Information about how they were recorded/coded are presented in the supplement.

We also included diagnoses of proinflammatory diseases ^45^ recorded across hospital inpatient records in either the primary or secondary position from 2006 onwards. For ease, the diseases we used will be referred to by the following names: arthritis, Crohn disease, systemic lupus erythematosus (SLE), Multiple Sclerosis (MS), chronic sinusitis, type two diabetes (T2D), hepatitis C, eczema and asthma. A detailed description of each disease is presented in the supplement.

We further grouped conditions into three clusters: atopy (asthma and eczema), chronic infection (hepatitis C and chronic sinusitis) and autoimmune disease (arthritis, T2D, SLE, MS, and Crohn disease). We created a count variable based on the number of conditions participants had within each disease cluster. Details of which cluster each disease is part of is presented in Supplementary Table 1.

### Statistical Analysis

GlycA was normally distributed, but hsCRP values were not, therefore both biomarkers were log-transformed (supplementary Figures 3-4 respectively). Z-scores were calculated so results were comparable.

We used ALSPAC clinic data at offspring mean ages 15y, 18y and 24y, and maternal mean ages 47y and 50yto investigate: 1) Short-term (weeks) correlations for GlycA and hsCRP using QC data; 2) Long-term (years) correlations for GlycA and hsCRP; and 3) Cross-sectional correlations between these markers at each clinic. We only had one QC measures of hsCRP (at the 24y clinic) which meant just one short-term correlation for CRP could be estimated. When investigating the mothers’ short term correlations using QC data, we used the clinic combined GlycA variable. We report only cross-sectional correlations between GlycA and hsCRP in UKB.

We examined associations between potential determinants of inflammation, GlycA and hsCRP using univariable and multivariable regression in ALSPAC and UKB. We also used UKB to investigate the association between inflammatory diseases and GlycA and CRP. Given that ALSPAC is a young/healthy cohort, there would be too few cases for a correctly powered analysis.

We investigated three models: a univariable analysis (model 1), models adjusting for age and sex (model 2; when age/sex were the exposure we adjusted for the other variable only), and models adjusting for other available determinants (model 3). A further description of the covariates included in model 3 is presented in the supplement.

## Results

Median GlycA and hsCRP levels, their range at different ages and their percentage coefficient of variance at each time point for ALSPAC and UKB are presented in Table 1 and 2 respectively. Characteristics of the main cohorts using ALSPAC offspring data and mother data are presented in Supplementary table 1. Characteristics of the UKB participants are presented in supplementary table 2.

**Table 1:** Means and standard deviations (SD) of GlycA and CRP at different timepoints in the ALSPAC offspring and mothers.

| Offspring | 15 y |  |  |  | 18y |  |  |  | 24y |  |  |  |
| --- | --- | --- | --- | --- | --- | --- | --- | --- | --- | --- | --- | --- |
|  | N | Median (IQR) | Range | % coefficient of variance | N | Median (IQR) | Range | % coefficient of variance | N | Median (IQR) | Range | % coefficient of variance |
| GlycA (mmol/L) | 3294 | 1.19 (1.12-1.28) | 0.87-1.90 | 10.62 | 3108 | 1.20 (1.12-1.29) | 0.90-1.93 | 11.14 | 3129 | 1.21 (1.11-1.33) | 0.84-2.25 | 13.87 |
| CRP (mmol/l) | 3374 | 0.01 (0.005-0.19) | 0.002-1.58 | 301.59 | 3189 | 0.01 ((0.006-0.03) | 0.0004-3.82 | 310.47 | 2958 | 0.02 (0.01-0.05) | 0.002-4.89 | 287.76 |
| Mothers | 48y |  |  |  | 50y |  |  |  | Combined group (47.57y) |  |  |  |
|  | N | Median (IQR) | Range | % coefficient of variance | N | Median (IQR) | Range | % coefficient of variance | N | Median (IQR) | Range | % coefficient of variance |
| GlycA (mmol/L) (SD) | 4243 | 1.22 (1.13-1.33) | 0.86-2.21 | 13.12 | 2709 | 1.23 (1.13-1.34) | 0.87-2.24 | 13.34 | 4453 | 1.22 (1.13-1.33) | 0.85-2.21 | 13.20 |
| CRP (mmol/l) (SD) | 4446 | 0.02 (0.01-0.05) | 0.001-1.32 | 168.94 | 2584 | 0.02 (0.01-0.05) | 0.004-1.66 | 184.85 | 4615 | 0.22 (0.1-0.5) | 0.001-1.32 | 246.48 |

**Table 2:**
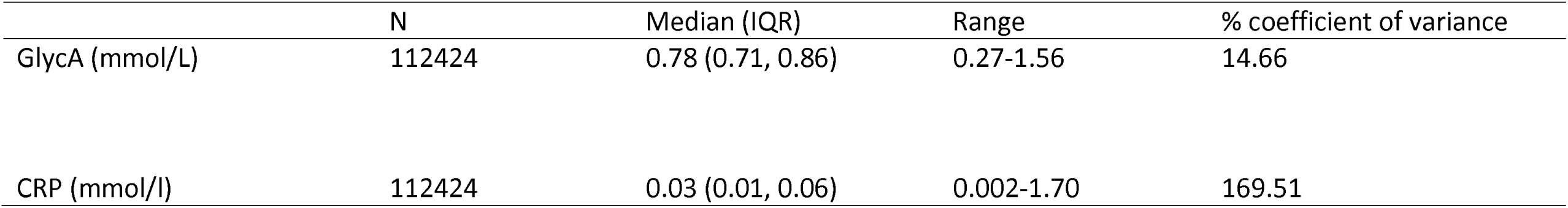
Means and standard deviations (SD) of GlycA and CRP at different timepoints in UKB (mean age = 48y)

### Correlations for GlycA and hsCRP

Short-term correlations of GlycA are presented in Figure 1, Panel A. There were moderate-to-strong correlations between measures of GlycA taken 5-6 weeks apart at ages 15y (r=0.75 [95% CI=0.56, 0.94], 18y (r=0.74 [0.64, 0.85], 24y (r=0.74 [0.51, 0.98] and 48y (r=0.82 [0.76, 0.86]. hsCRP had a strong short-term correlation (r=0.77 [0.59, 0.95] at age 24y.

**Figure 1:**
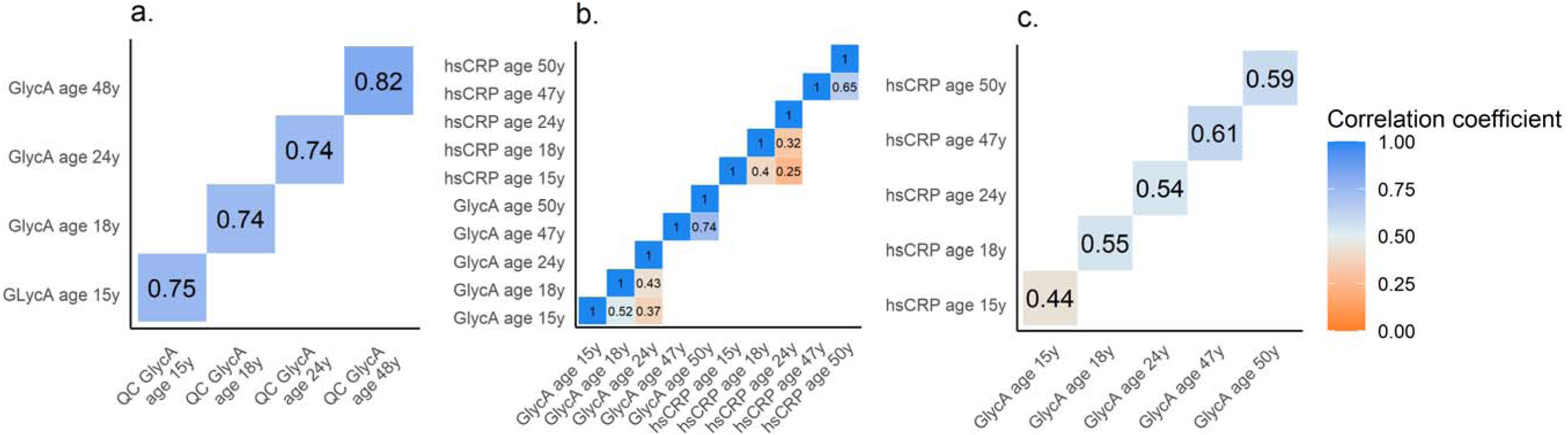
Correlation plots using the ALSPAC cohort (both mothers and offspring) a. Short-term correlations (5-6 weeks) between logged GlycA at clinic and logged GlycA QC data. b. Long-term correlations of logged GlycA and logged hsCRP at different timepoints across the life course. c: Cross-sectional correlations between logged GlycA and logged hsCRP across the life course.

GlycA had moderate correlations between measures taken years apart in adolescence/early-adulthood (between 15y and 18y: r=0.52 [0.47, 0.56]; between 15y and 24y: r=0.37 [0.31, 0.44]; between 18y and 24y: r=0.43 [0.37, 0.49] and strong correlations in mid-adulthood (between 47y and 50y: r=0.74 [0.72, 0.76]. These were larger than equivalent correlations of hsCRP (between 15y and 18y: r=0.40 [0.35, 0.46] between 15y and 24y: r=0.25 [0.18, 0.32]; between 18y and 24y: r=0.32 [0.25, 0.38]; between 47y and 50y: r=0.65 [0.62, 0.68]. Figure 1, Panel B shows the long-term correlations for GlycA and hsCRP (separately).

Concurrently measured GlycA and hsCRP levels were moderately correlated at all ages: 15y (r=0.44 [0.40, 0.48], 18y (r=0.55 [0.51, 0.59], 24y (r=0.54 [0.50, 0.59], 47y (r=0.61 [0.59, 0.63] and 50y (r=0.59 [0.56, 0.61] are presented in Figure 1, Panel C. In UKB, the cross-sectional correlation between GlycA and hsCRP (r=0.52 [0.52, 0.53] was similar to correlations when using the ALSPAC data.

As a sensitivity analysis, we re-estimated correlations excluding all individuals who reported experiencing an infection at any clinic. This was to mitigate the short-term effects an infection may have; results were comparable to the primary analysis and are presented in the supplement.

We also present Bland-Altman plots for each of the correlations in the ALSPAC offspring and the mothers to indicate the agreement of the measures and these are present in supplementary figure 5 and 6 respectively.

### Associations with potential determinants of inflammation

#### ALSPAC offspring

Estimates from both univariable and multivariable models are presented in Table 3 for GlycA and Table 4 for hsCRP. In models including all available potential determinants (model 3; Figure 2), BMI and having a recent infection were positively associated with GlycA and hsCRP at the time points they were recorded. Additionally, in model 3, GlycA was higher in females than males at ages 15y and 18y, but attenuated to the null at 24y. Conversely, mean hsCRP was higher in males at ages 15y but was then higher in females in later ages. Lower social class was positively associated with hsCRP at 15y only and GlycA at 25y only. Mother’s educational qualification, smoking frequency and alcohol intake were not associated with GlycA/hsCRP at any time point.

**Figure 2:**
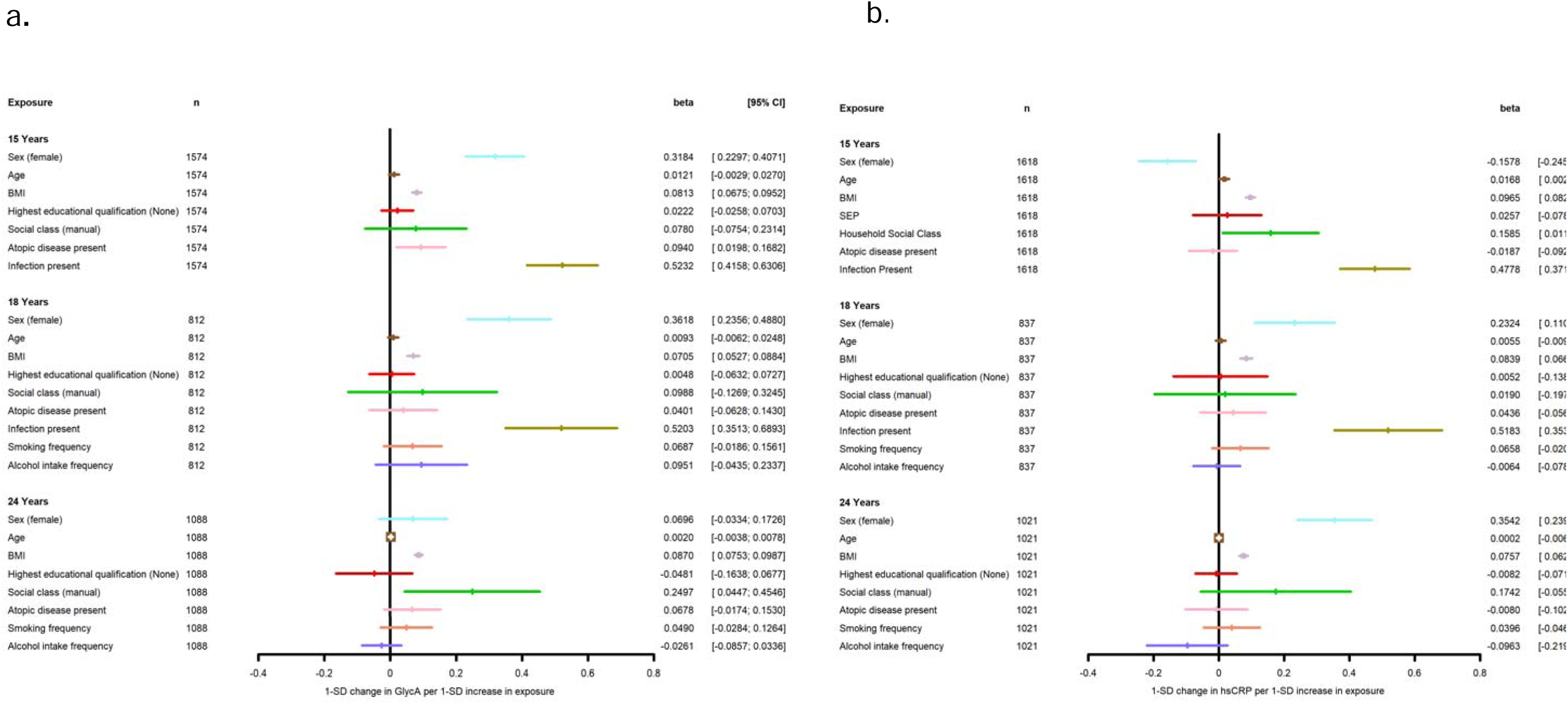
Associations between known determinants of inflammation and the biomarkers hsCRP and GlycA in the ALSPAC offspring at ages 15y, 18y and 24y. SEP = Mother holds Degree vs No degree, Household Social Class = Parent’s manual vs Non-manual occupation, Atopy = Asthma and eczema, asthma or eczema or neither, Smoking frequency= none, less than once a week, more than once a week, Drinking frequency= none, less than once a week, more than once a week. Both biomarkers are log-transformed

**Table 3:**
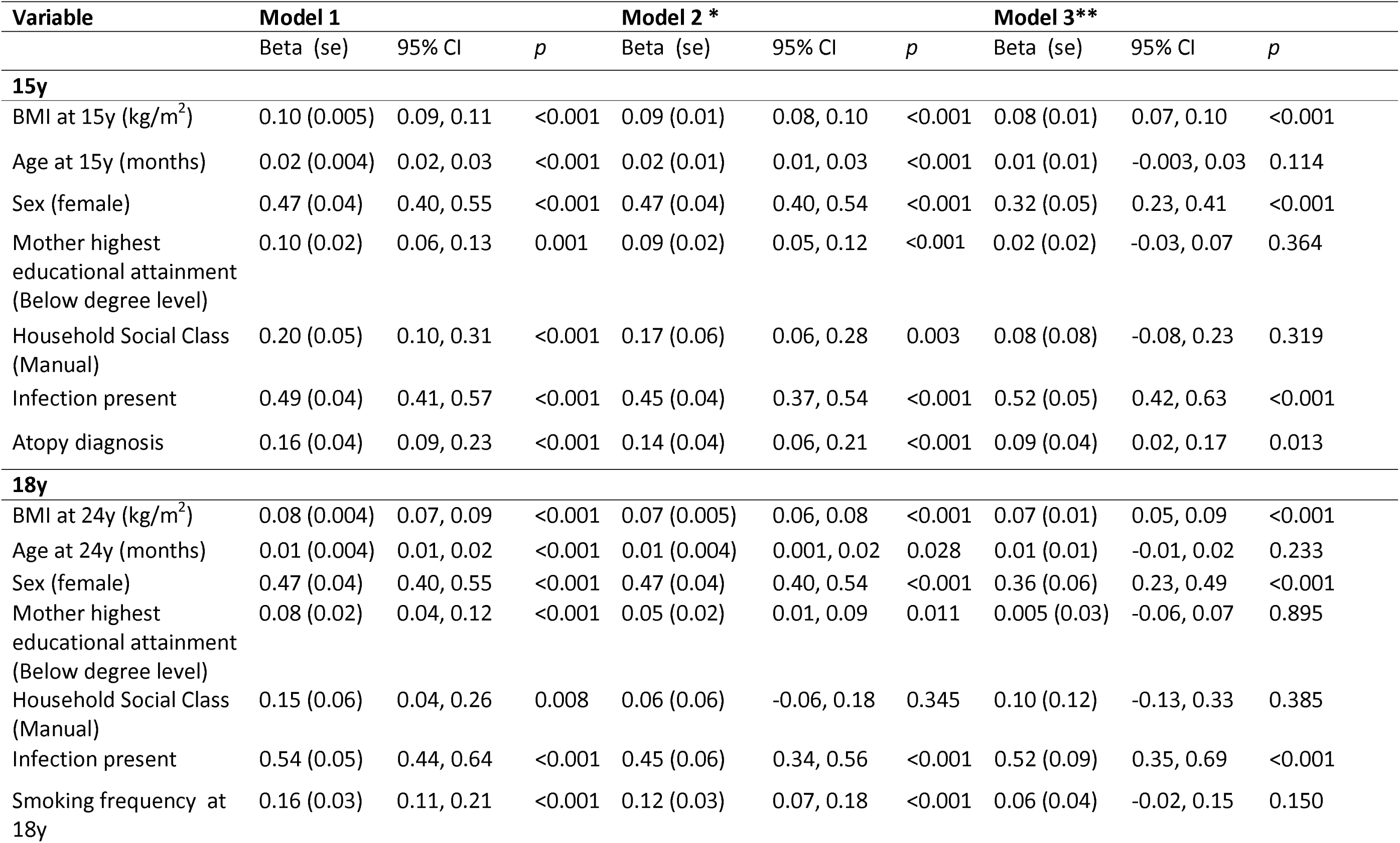
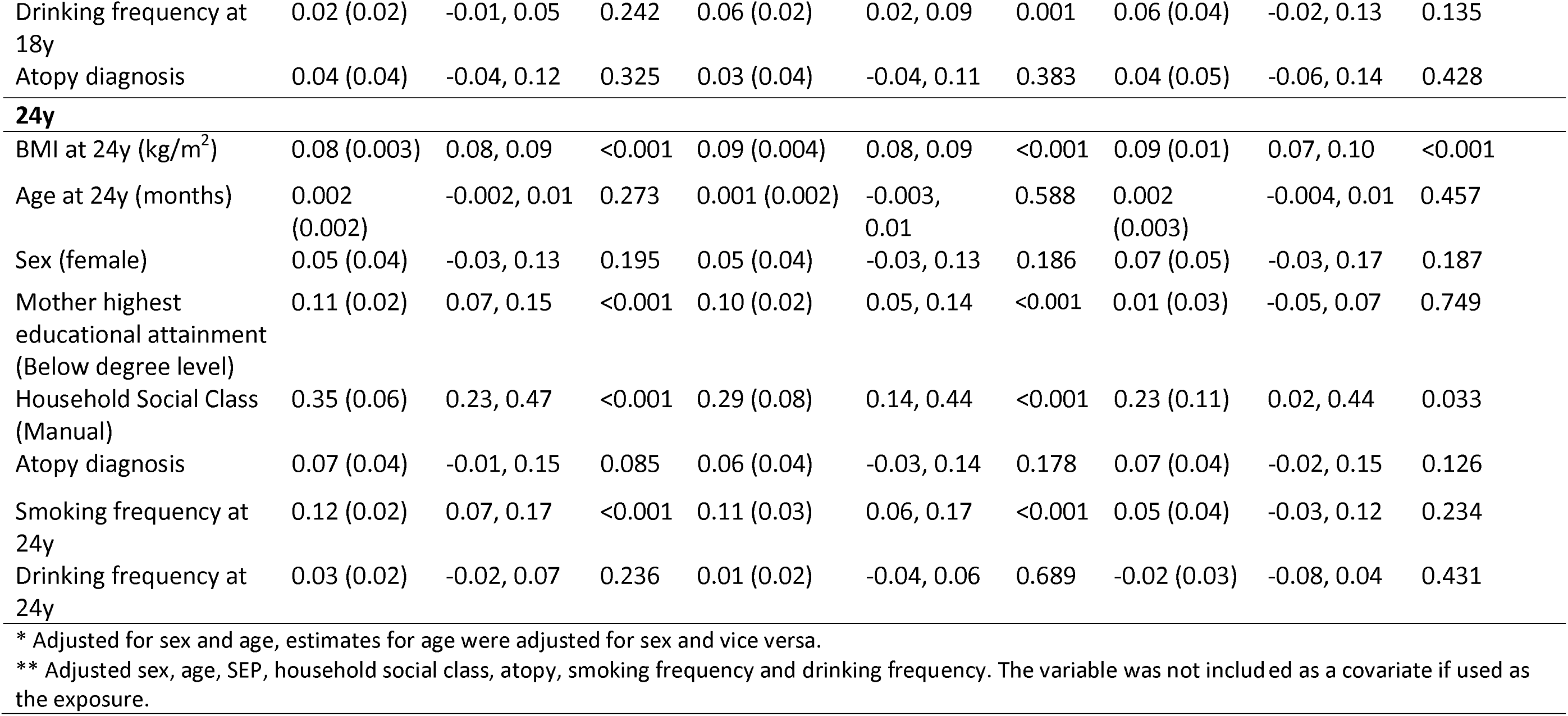
Cross-sectional associations between key determinants of inflammation and GlycA at 15y, 18y and 24y.

| Variable | Model 1 |  |  | Model 2 * |  |  | Model 3 ** |  |  |
| --- | --- | --- | --- | --- | --- | --- | --- | --- | --- |
|  | Beta (se) | 95% CI | *p* | Beta (se) | 95% CI | *p* | Beta (se) | 95% CI | *p* |
| **15y** |  |  |  |  |  |  |  |  |  |
| BMI at 15y (kg/m<sup>2</sup>) | 0.10 (0.005) | 0.09, 0.11 | <0.001 | 0.09 (0.01) | 0.08, 0.10 | <0.001 | 0.08 (0.01) | 0.07, 0.10 | <0.001 |
| Age at 15y (months) | 0.02 (0.004) | 0.02, 0.03 | <0.001 | 0.02 (0.01) | 0.01, 0.03 | <0.001 | 0.01 (0.01) | -0.003, 0.03 | 0.114 |
| Sex (female) | 0.47 (0.04) | 0.40, 0.55 | <0.001 | 0.47 (0.04) | 0.40, 0.54 | <0.001 | 0.32 (0.05) | 0.23, 0.41 | <0.001 |
| Mother highest educational attainment (Below degree level) | 0.10 (0.02) | 0.06, 0.13 | 0.001 | 0.09 (0.02) | 0.05, 0.12 | <0.001 | 0.02 (0.02) | -0.03, 0.07 | 0.364 |
| Household Social Class (Manual) | 0.20 (0.05) | 0.10, 0.31 | <0.001 | 0.17 (0.06) | 0.06, 0.28 | 0.003 | 0.08 (0.08) | -0.08, 0.23 | 0.319 |
| Infection present | 0.49 (0.04) | 0.41, 0.57 | <0.001 | 0.45 (0.04) | 0.37, 0.54 | <0.001 | 0.52 (0.05) | 0.42, 0.63 | <0.001 |
| Atopy diagnosis | 0.16 (0.04) | 0.09, 0.23 | <0.001 | 0.14 (0.04) | 0.06, 0.21 | <0.001 | 0.09 (0.04) | 0.02, 0.17 | 0.013 |
| **18y** |  |  |  |  |  |  |  |  |  |
| BMI at 24y (kg/m<sup>2</sup>) | 0.08 (0.004) | 0.07, 0.09 | <0.001 | 0.07 (0.005) | 0.06, 0.08 | <0.001 | 0.07 (0.01) | 0.05, 0.09 | <0.001 |
| Age at 24y (months) | 0.01 (0.004) | 0.01, 0.02 | <0.001 | 0.01 (0.004) | 0.001, 0.02 | 0.028 | 0.01 (0.01) | -0.01, 0.02 | 0.233 |
| Sex (female) | 0.47 (0.04) | 0.40, 0.55 | <0.001 | 0.47 (0.04) | 0.40, 0.54 | <0.001 | 0.36 (0.06) | 0.23, 0.49 | <0.001 |
| Mother highest educational attainment (Below degree level) | 0.08 (0.02) | 0.04, 0.12 | <0.001 | 0.05 (0.02) | 0.01, 0.09 | 0.011 | 0.005 (0.03) | -0.06, 0.07 | 0.895 |
| Household Social Class (Manual) | 0.15 (0.06) | 0.04, 0.26 | 0.008 | 0.06 (0.06) | -0.06, 0.18 | 0.345 | 0.10 (0.12) | -0.13, 0.33 | 0.385 |
| Infection present | 0.54 (0.05) | 0.44, 0.64 | <0.001 | 0.45 (0.06) | 0.34, 0.56 | <0.001 | 0.52 (0.09) | 0.35, 0.69 | <0.001 |
| Smoking frequency at 18y | 0.16 (0.03) | 0.11, 0.21 | <0.001 | 0.12 (0.03) | 0.07, 0.18 | <0.001 | 0.06 (0.04) | -0.02, 0.15 | 0.150 |
| Drinking frequency at 18y | 0.02 (0.02) | -0.01, 0.05 | 0.242 | 0.06 (0.02) | 0.02, 0.09 | 0.001 | 0.06 (0.04) | -0.02, 0.13 | 0.135 |
| Atopy diagnosis | 0.04 (0.04) | -0.04, 0.12 | 0.325 | 0.03 (0.04) | -0.04, 0.11 | 0.383 | 0.04 (0.05) | -0.06, 0.14 | 0.428 |
| 24y |  |  |  |  |  |  |  |  |  |
| BMI at 24y (kg/m<sup>2</sup>) | 0.08 (0.003) | 0.08, 0.09 | <0.001 | 0.09 (0.004) | 0.08, 0.09 | <0.001 | 0.09 (0.01) | 0.07, 0.10 | <0.001 |
| Age at 24y (months) | 0.002 (0.002) | -0.002, 0.01 | 0.273 | 0.001 (0.002) | -0.003, 0.01 | 0.588 | 0.002 (0.003) | -0.004, 0.01 | 0.457 |
| Sex (female) | 0.05 (0.04) | -0.03, 0.13 | 0.195 | 0.05 (0.04) | -0.03, 0.13 | 0.186 | 0.07 (0.05) | -0.03, 0.17 | 0.187 |
| Mother highest educational attainment (Below degree level) | 0.11 (0.02) | 0.07, 0.15 | <0.001 | 0.10 (0.02) | 0.05, 0.14 | <0.001 | 0.01 (0.03) | -0.05, 0.07 | 0.749 |
| Household Social Class (Manual) | 0.35 (0.06) | 0.23, 0.47 | <0.001 | 0.29 (0.08) | 0.14, 0.44 | <0.001 | 0.23 (0.11) | 0.02, 0.44 | 0.033 |
| Atopy diagnosis | 0.07 (0.04) | -0.01, 0.15 | 0.085 | 0.06 (0.04) | -0.03, 0.14 | 0.178 | 0.07 (0.04) | -0.02, 0.15 | 0.126 |
| Smoking frequency at 24y | 0.12 (0.02) | 0.07, 0.17 | <0.001 | 0.11 (0.03) | 0.06, 0.17 | <0.001 | 0.05 (0.04) | -0.03, 0.12 | 0.234 |
| Drinking frequency at 24y | 0.03 (0.02) | -0.02, 0.07 | 0.236 | 0.01 (0.02) | -0.04, 0.06 | 0.689 | -0.02 (0.03) | -0.08, 0.04 | 0.431 |

\* Adjusted for sex and age, estimates for age were adjusted for sex and vice versa.
\*\* Adjusted sex, age, SEP, household social class, atopy, smoking frequency and drinking frequency. The variable was not included as a covariate if used as the exposure.

**Table 4:**
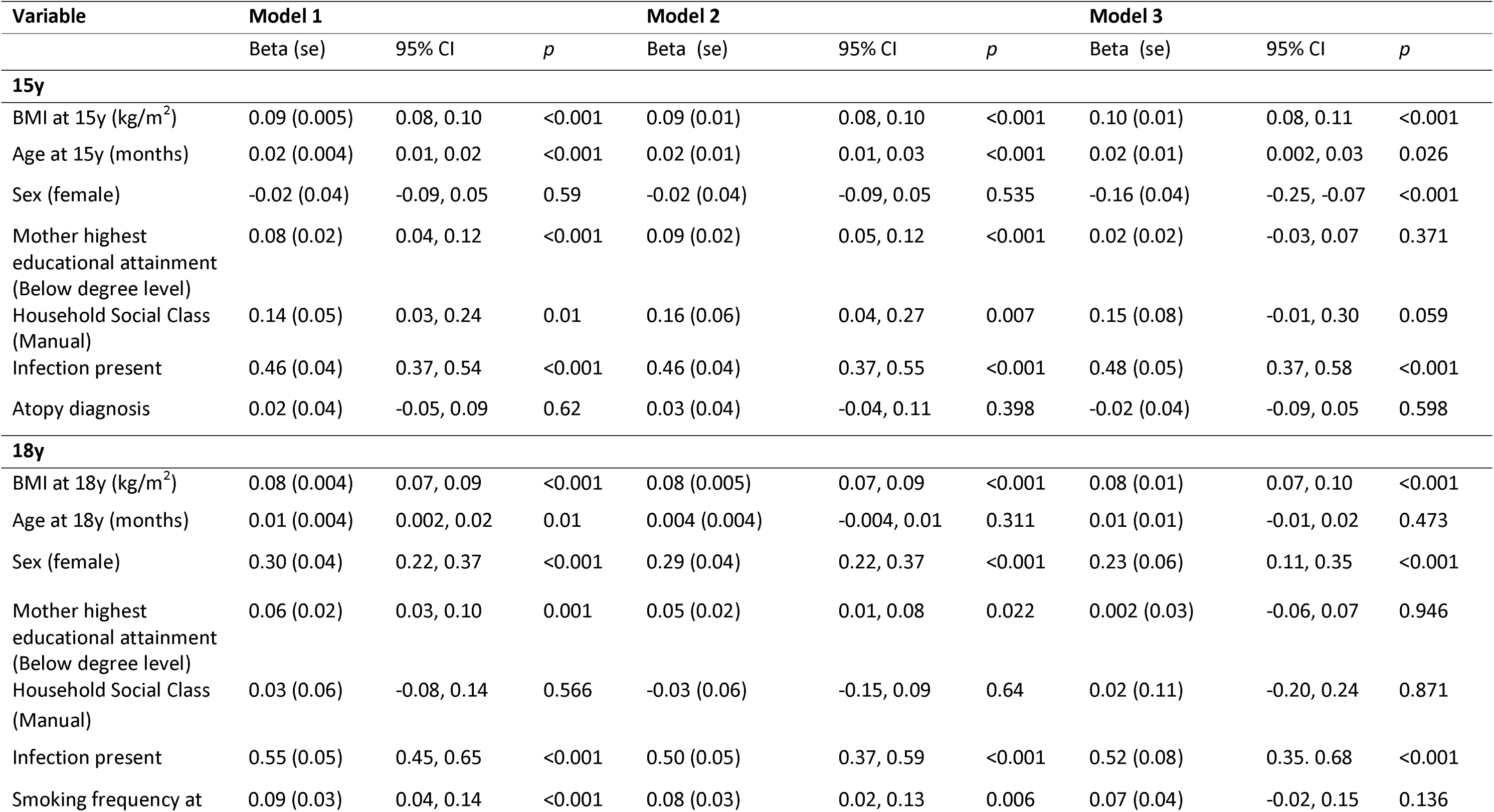
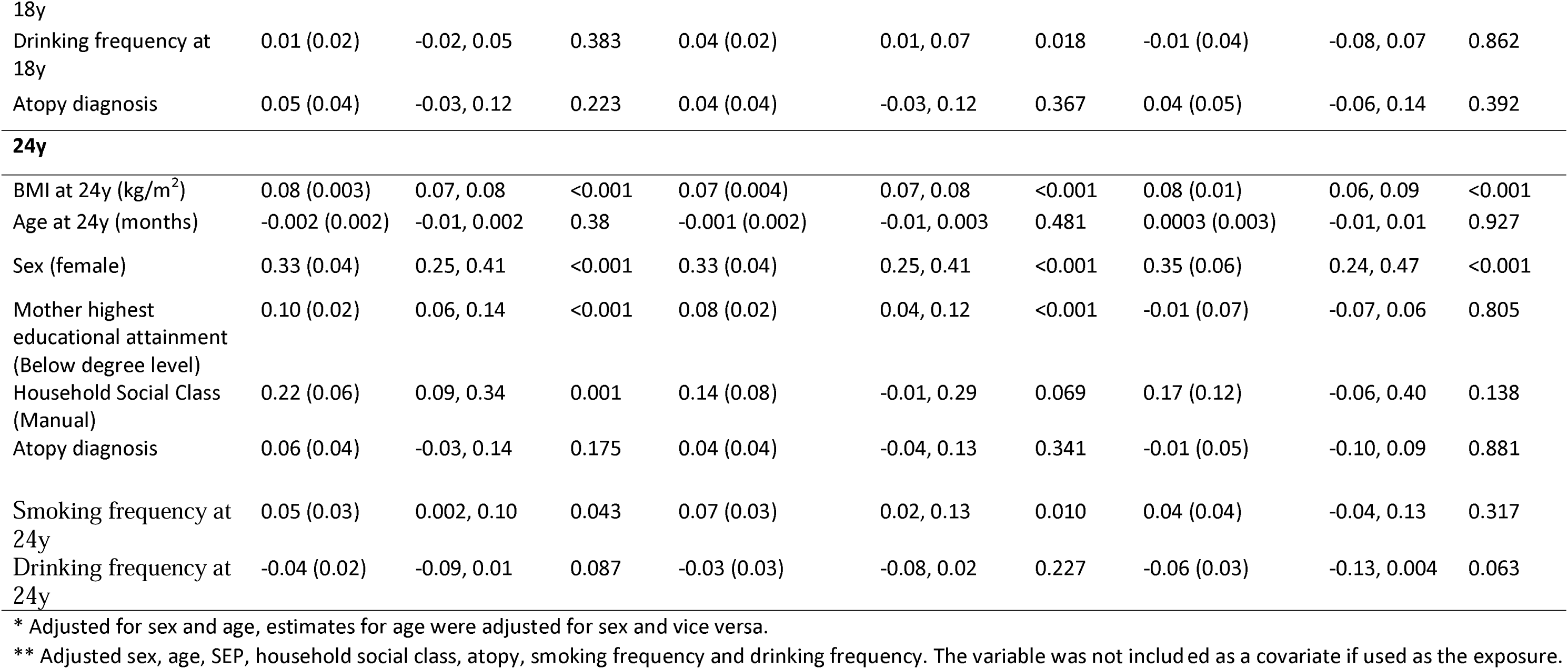
Cross-sectional associations between key determinants of inflammation and CRP at 15y, 18y and 24y.

#### ALSPAC mothers

Estimates from the univariable and multivariable models are presented in Table 5. In model 3, BMI, age and lower academic achievement were positively associated with GlycA and hsCRP. Manual household social class was positively associated with GlycA only. The only inflammation-related factor which did not show an association with either GlycA or hsCRP was an atopy diagnosis (Figure 3).

**Figure 3:**
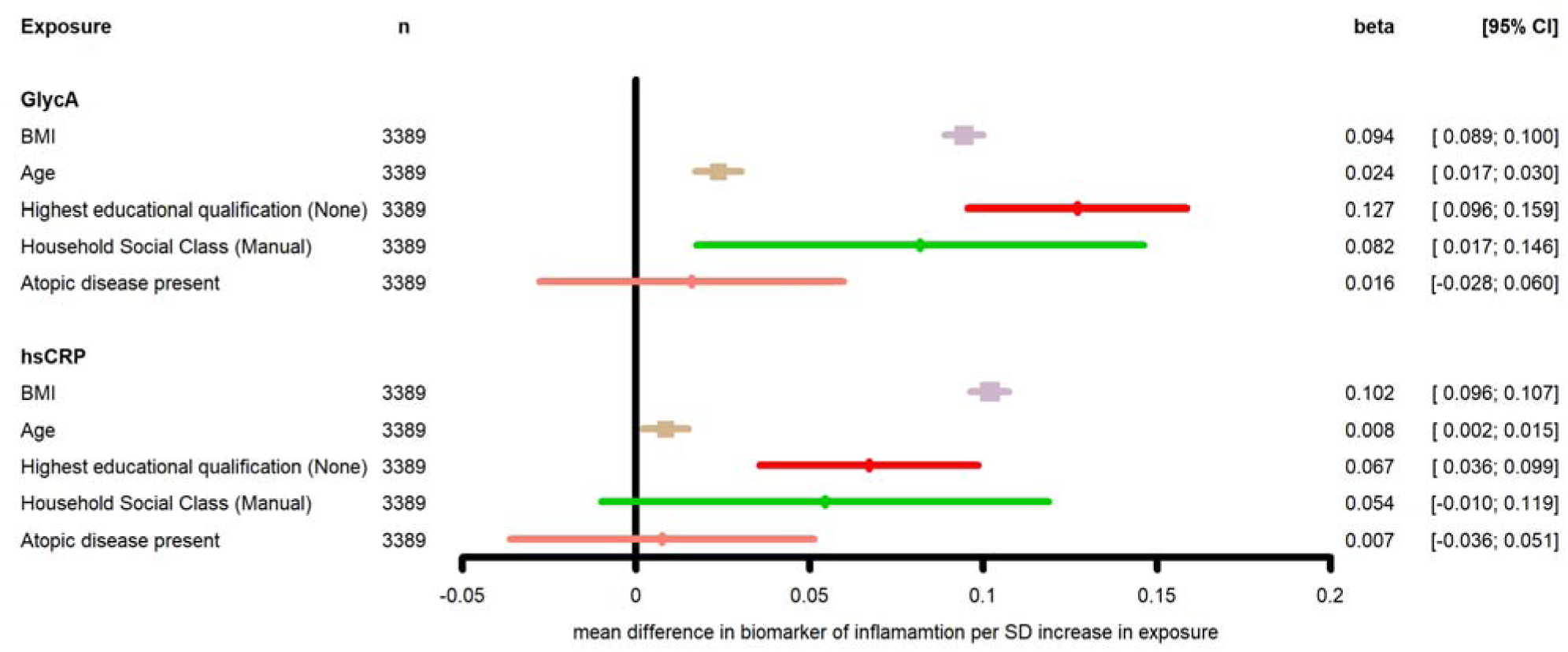
Association between known determinants of inflammation and GlycA and hsCRP in ALSPAC mothers

**Table 5:** Cross-sectional associations between key determinants of inflammation and inflammatory biomarkers in the mothers cohort at mean age 47.57y.

| Variable | Model 1 |  |  | Model 2 |  |  | Model 3 |  |  |
| --- | --- | --- | --- | --- | --- | --- | --- | --- | --- |
|  | Beta (se) | 95% CI | *p* | Beta (se) | 95% CI | *p* | Beta (se) | 95% CI | *p* |
| **GlycA** |  |  |  |  |  |  |  |  |  |
| BMI (kg/m<sup>2</sup>) | 0.10 (0.002) | 0.09, 0.10 | <0.001 | 0.10 (0.002) | 0.09, 0.10 | <0.001 | 0.09 (0.003) | 0.09, 0.10 | <0.001 |
| Age (months) | 0.004 (0.003) | -0.002, 0.01 | 0.176 | - | - | - | 0.02 (0.003) | 0.02, 0.03 | <0.001 |
| SEP (below degree level) | 0.19 (0.02) | 0.15, 0.22 | <0.001 | 0.21 (0.02) | 0.18, 0.24 | <0.001 | 0.13 (0.02) | 0.10, 0.16 | <0.001 |
| Grandparents Social Class (manual) | 0.23 (0.04) | 0.16, 0.30 | <0.001 | 0.24 (0.04) | 0.17, 0.31 | <0.001 | 0.08 (0.03) | 0.02, 0.15 | 0.013 |
| Atopic diagnosis | 0.04 (0.02) | -0.01, 0.08 | 0.133 | 0.04 (0.02) | -0.01, 0.09 | 0.115 | 0.02 (0.02) | -0.03, 0.06 | 0.474 |
| **CRP** |  |  |  |  |  |  |  |  |  |
| BMI (kg/m<sup>2</sup>) | 0.10 (0.002) | 0.10, 0.11 | <0.001 | 0.10 (0.002) | 0.10, 0.11 | <0.001 | 0.10 (0.003) | 0.10, 0.11 | <0.001 |
| Age (months) | -0.004 (0.003) | -0.01, 0.002 | 0.278 | - | - | - | 0.01 (0.003) | 0.002, 0.02 | 0.013 |
| SEP (below degree level) | 0.15 (0.02) | 0.12, 0.18 | <0.001 | 0.16 (0.02) | 0.12, 0.19 | <0.001 | 0.07 (0.02) | 0.04, 0.10 | <0.001 |
| Grandparents Social Class (manual) | 0.18 (0.04) | 0.11, 0.25 | <0.001 | 0.18 (0.04) | 0.11, 0.25 | <0.001 | 0.05 (0.03) | -0.01, 0.12 | 0.097 |
| Atopic diagnosis | 0.04 (0.02) | -0.01, 0.09 | 0.155 | 0.04 (0.02) | -0.01, 0.09 | 0.137 | 0.01 (0.02) | -0.04, 0.05 | 0.738 |

\* Adjusted age, estimates for age were left as in Model 1
\*\* Adjusted for age, SEP, grandparents household social class and atopy. The variable was not included as a covariate if used as the exposure.

#### UKB

Estimates from the univariable and multivariable models for associations between key characteristics and behaviours and both GlycA and hsCRP are presented in Table 6. As in ALSPAC, BMI was positively associated with both GlycA and hsCRP in the unadjusted and adjusted models. Females had lower levels of inflammation (measured by GlycA and hsCRP) in the adjusted and unadjusted models. Being a frequent drinker was negatively associated with levels of GlycA in adjusted and unadjusted models. Additionally, we found that lower SEP and being a frequent smoker were associated with higher inflammation measured by GlycA and hsCRP in all three models. Results from model 3 are presented in Figure 4. As a sensitivity analysis, we re-ran the estimates excluded all individuals that had a diagnosis of the included inflammatory-related diseases. Results are presented in supplementary Table 4 and were comparable to the unrestricted dataset.

**Figure 4:**
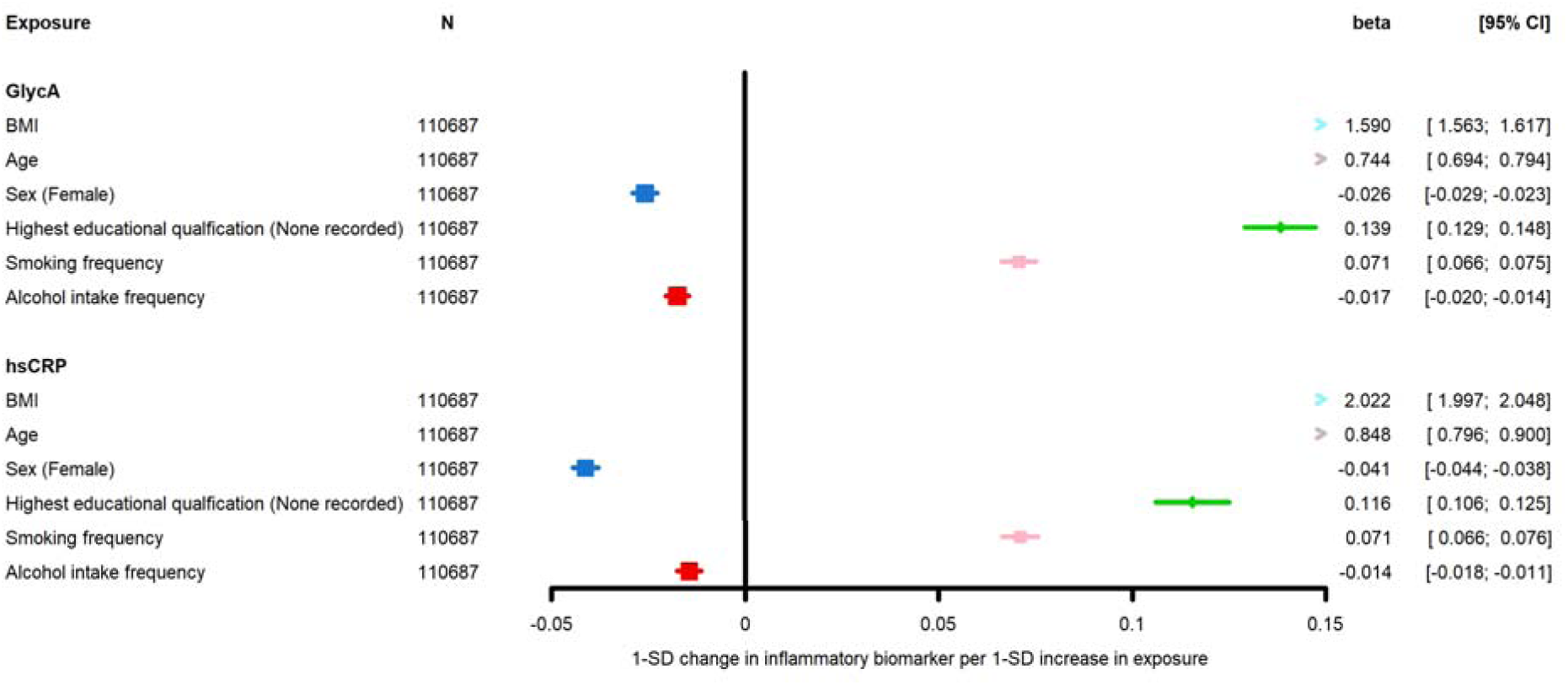
Association between known determinants of inflammation and GlycA and hsCRP in UKB

**Table 6:** Cross-sectional associations between key determinants of inflammation and GlycA in UKB.

| Variable | Model 1 |  |  | Model 2 * |  |  | Model 3 ** (N=91591) |  |  |
| --- | --- | --- | --- | --- | --- | --- | --- | --- | --- |
|  | Beta (se) | 95% CI | p | Beta (se) | 95% CI | p | Beta (se) | 95% CI | p |
| **GlycA** |  |  |  |  |  |  |  |  |  |
| BMI at (kg/m<sup>2</sup>) | 1.64 (0.01) | 1.61, 1.67 | <0.001 | 1.64 (0.01) | 1.62, 1.67 | <0.001 | 1.59 (0.01) | 1.56, 1.62 | <0.001 |
| Age (months) | 0.95 (0.02) | 0.90, 0.99 | <0.001 | 0.95 (0.02) | 0.90, 1.00 | <0.001 | 0.74 (0.03) | 0.69, 0.79 | <0.001 |
| Sex (female) | -0.01 (0.002) | -0.01, -0.003 | <0.001 | -0.01 (0.002) | -0.01, -0.004 | <0.001 | -0.03 (0.002) | -0.03, -0.02 | <0.001 |
| Highest educational attainment (Below degree level) | 0.24 (0.002) | 0.23, 0.25 | <0.001 | 0.21 (0.004) | 0.20, 0.22 | <0.001 | 0.14 (0.005) | 0.13, 0.15 | <0.001 |
| Smoking frequency | 0.07 (0.002) | 0.07, 0.074 | <0.001 | 0.07 (0.002) | 0.07, 0.07 | <0.001 | 0.07 (0.002) | 0.07, 0.09 | <0.001 |
| Drinking frequency | -0.03 (0.001) | -0.03, -0.02 | <0.001 | -0.02 (0.001) | -0.03, -0.02 | <0.001 | -0.02 (0.002) | -0.02, -0.01 | <0.001 |
| **CRP** |  |  |  |  |  |  |  |  |  |
| BMI (kg/m<sup>2</sup>) | 2.05 (0.01) | 2.02, 2.07 | <0.001 | 2.06 (0.01) | 2.04, 2.09 | <0.001 | 2.02 (0.01) | 1.91, 2.05 | <0.001 |
| Age (months) | 0.98 (0.02) | 0.93, 1.02 | <0.001 | 0.98 (0.02) | 0.94, 1.03 | <0.001 | 0.85 (0.03) | 0.82, 0.90 | <0.001 |
| Sex (female) | -0.01 (0.002) | -0.02, -0.01 | <0.001 | -0.01 (0.001) | -0.02, -0.01 | <0.001 | -0.04 (0.002) | -0.043, -0.04 | <0.001 |
| Highest educational attainment (Below degree level) | 0.23 (0.004) | 0.22, 0.24 | <0.001 | 0.20 (0.004) | 0.20, 0.21 | <0.001 | 0.12 (0.005) | 0.11, 0.13 | <0.001 |
| Smoking frequency | 0.06 (0.002) | 0.06, 0.07 | <0.001 | 0.06 (0.002) | 0.06, 0.07 | <0.001 | 0.07 (0.02) | 0.07, 0.08 | <0.001 |
| Drinking frequency | -0.02 (0.001) | -0.03, 0.02 | <0.001 | -0.02 (0.001) | -0.03, -0.02 | <0.001 | -0.01 (0.01) | -0.02, -0.01 | <0.001 |

\* Adjusted for sex and age, estimates for age were adjusted for sex and vice versa.
\*\* Adjusted sex, age, SEP, smoking frequency and drinking frequency. The variable was not included as a covariate if used as the exposure.

Estimates from the univariable and multivariable models examining association between known inflammatory diseases with GlycA and CRP are presented in Table 7. Multiple inflammation-related conditions were associated with both GlycA and hsCRP in a similar pattern. In model 3, arthritis, asthma, MS, chronic sinusitis and Crohn’s disease were associated with both higher GlycA and hsCRP; T2D was positively associated with GlycA only; and eczema was associated with hsCRP only. In all three models, neither SLE or hepatitis C were associated with GlycA or hsCRP.

**Table 7:**
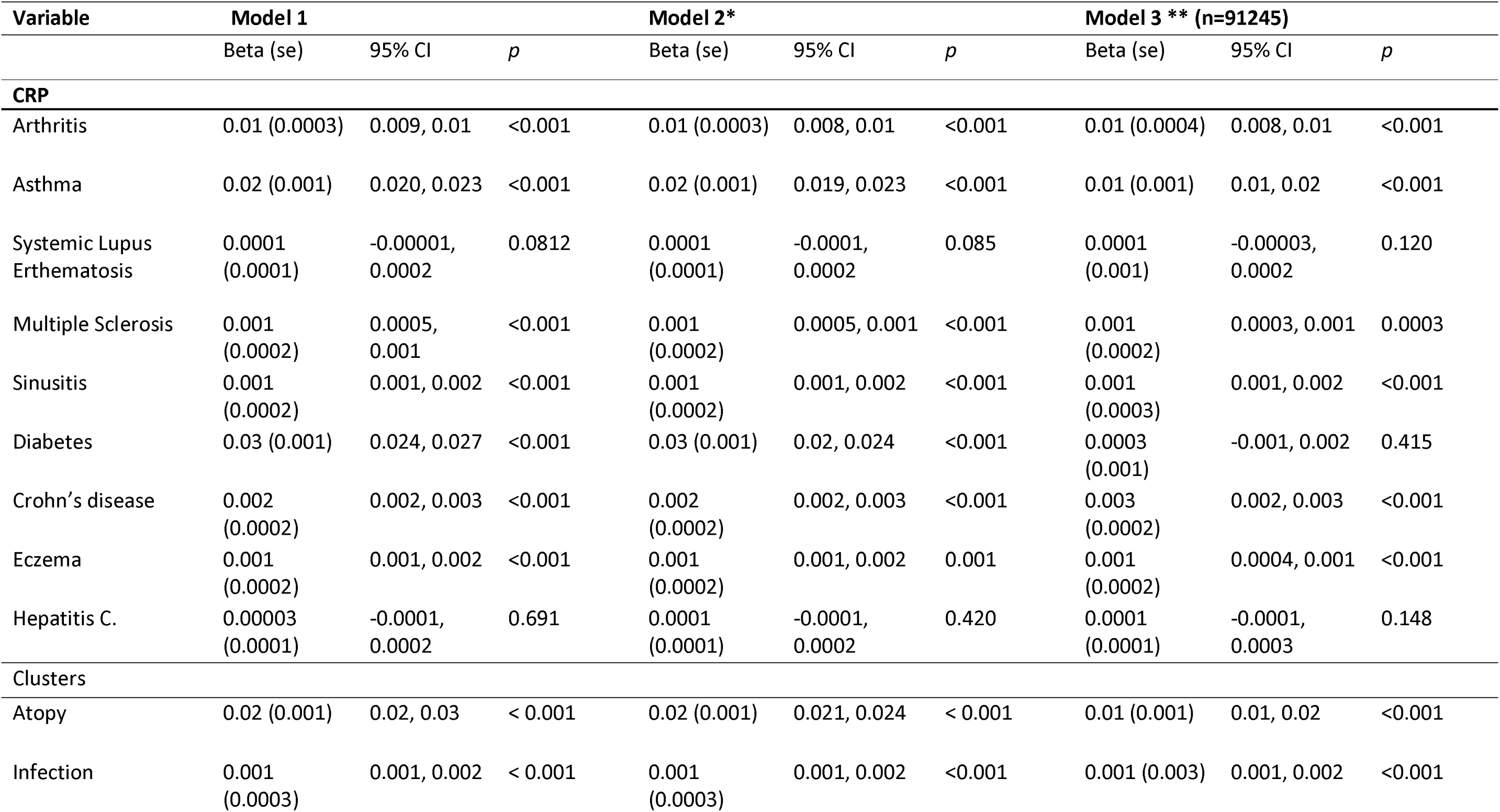
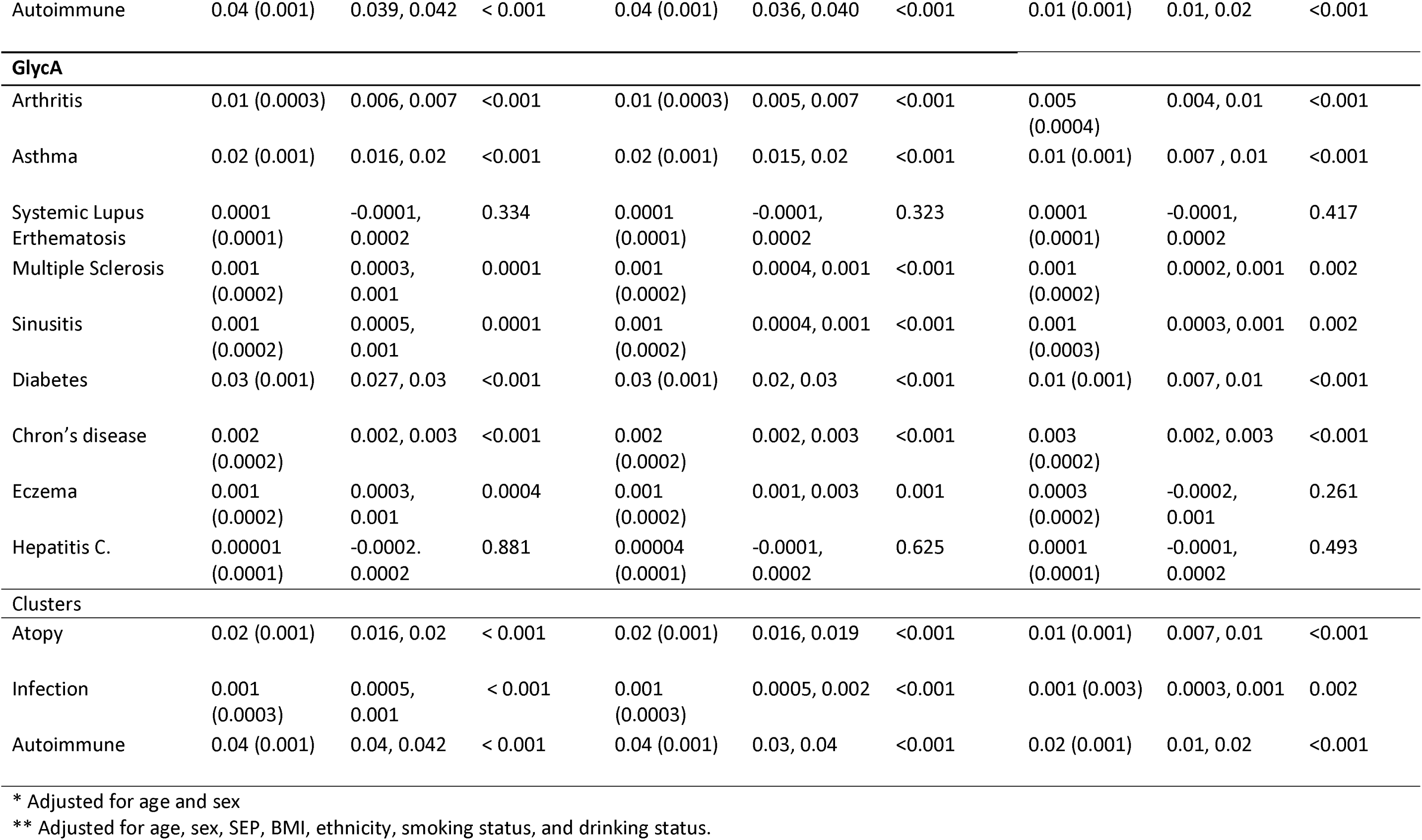
Cross-sectional associations between inflammation-related diseases and GlycA and hsCRP in UKB.

The atopy cluster and the autoimmune cluster were both positively associated with GlycA and hsCRP in all three models. The infection cluster was positively associated with hsCRP in all three models and with GlycA in model 1 and 2. However, in model 3, the confidence interval for GlycA crossed the null value (Figure 5).

**Figure 5:**
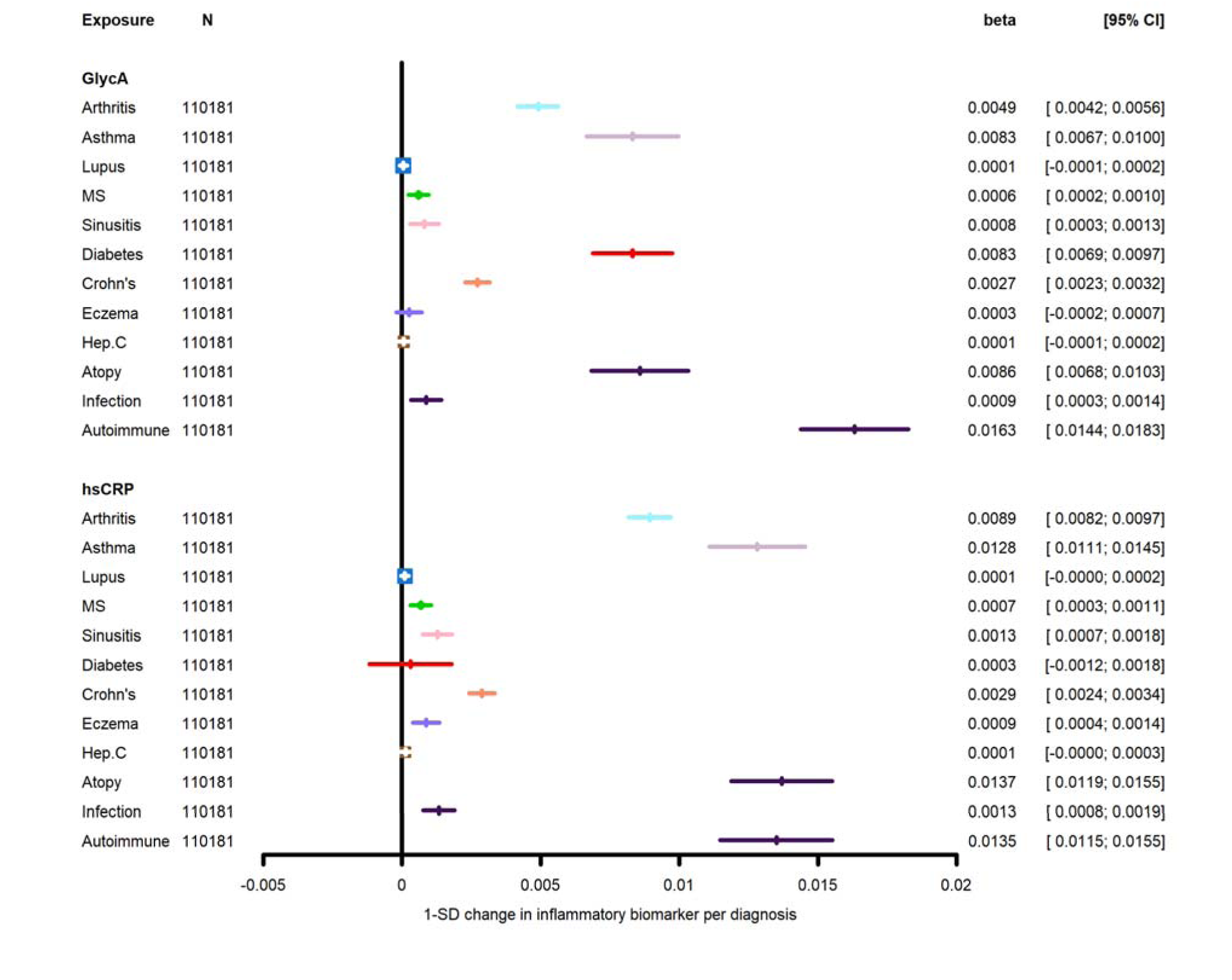
Associations between known inflammatory diseases and the biomarkers CRP and GlycA in the UKB Both biomarkers are log-transformed

Further description of associations in models 1 and 2 using the ALSPAC and the UKB data are presented in the supplement.

## Discussion

We found that the short-term correlation of hsCRP was similar in magnitude to the short-term GlycA correlations and GlycA and hsCRP were moderately correlated with each other throughout the life-course. Long-term intra-biomarker correlations for GlycA were slightly stronger than those for hsCRP. The findings were similar in magnitude to the correlation estimated by a study by Ritchie et al. which used 3 repeated GlycA measures taken over the course of a decade (r=0.43) ^46^. GlycA also displayed a lower coefficient of variation compared to hsCRP using ALSPAC and UKB data which suggests that GlycA has lower variability around the mean compared to hsCRP. The limits of agreement are narrower for GlycA than for hsCRP shown by the Bland-Alman plots which suggests that the measures between timepoints are more stable for GlycA than for hsCRP.

We found that having an infection at the time of blood sampling was positively associated with GlycA and hsCRP. This is expected as GlycA is a composite marker of acute-phase proteins and therefore its signal will increase in amplitude in response to an acute inflammatory stimulus. Despite this, the long-term correlations suggest that GlycA has greater stability across the life-course compared to hsCRP, which is in line with previous literature ^47^. The difference in stability between GlycA and hsCRP may also underlie previous findings that GlycA remains associated with proinflammatory diseases such as CVD, after adjusting for hsCRP ^39^. This may be because GlycA and hsCRP reflect overlapping but different inflammatory pathways ^48^. By logging hsCRP, variation in hsCRP is reduced (see supplementary Figure 4) and our regression results became comparable.

There were differences between associations in ALSPAC compared to UKB. Alcohol intake was not associated with either biomarker when using ALSPAC data, but it was negatively associated with levels of GlycA and hsCRP when using UKB data. UKB participants are older than those in ALSPAC and therefore a higher number of individuals may have inflammatory-related diseases. “Healthy” individuals may be more likely to drink, but still have lower levels of inflammation compared to individuals who have a pro-inflammatory disease, but no longer drink. Although the negative association was maintained in our secondary analysis, which aimed to account for this effect, other diseases were not included in our model which could be driving this negative association.

The magnitude of association between the infection variable and hsCRP and GlycA in ALSPAC was larger than that in UKB, which is likely because in ALSPAC, this variable codes for infection within the preceding 3 weeks while in UKB the variable reflects a diagnosis of Hepatitis C or chronic sinusitis from 2006 onwards and which are chronic or recurrent infections, respectively. We also found a positive association between atopy and hsCRP and GlycA levels when using UKB data, but not in ALSPAC offspring or mothers. This may be because UKB classifies diseases based on hospitalisation only and therefore is likely to capture only the more severe cases. ALSPAC participants self-reported diagnoses of an atopic disease. Although, this can also include diagnoses made in a primary care setting, it is more likely to include less severe cases (mild eczema vs anaphylaxis) which contribute to the attenuation of an effect in the ALSPAC cohort. In addition, the sample size in the UKB analysis was approximately 20 times larger than that in the ALSPAC analysis and therefore there was more power to detect associations in UKB.

### Strengths and Limitations

We used two large population-based cohorts, covering a wide range of ages which allowed us to compare the short-term and long-term stability of both biomarkers and the performance of GlycA and hsCRP as biomarkers throughout the life-course.

We acknowledge some limitations. Short-term repeat hsCRP data were unavailable for most clinics, so it was only possible to assess short-term stability of hsCRP at 24yrs. Some variables such as smoking frequency and alcohol intake rely on self-report and not all variables (such as infection) were measured at every timepoint. Additionally, several of the diseases are characterised by acute flares in disease activity, but this information was not provided and so we could not differentiate whether levels of the biomarkers reflected chronic inflammation or acute flares in disease activity.

## Conclusion

In conclusion, we found that GlycA and hsCRP correlated moderately across the life-course, with some sex differences evident at different ages. The short-term GlycA correlations were comparable to the short-term hsCRP correlation, as were associations of pro-inflammatory factors and diseases with GlycA and hsCRP. Nevertheless, GlycA showed greater long-term stability compared to hsCRP.

## Data Availability

Data needed to evaluate the conclusions presented in this paper are provided in the manuscript and/or the Supplementary Materials. Additional ALSPAC data can be requested from the ALSPAC executive committee and reasonable requests from bona fide researchers will be approved. This research has been conducted using data from UK Biobank project ID:81499, a major biomedical database and can be provided by UKB (http://www.ukbiobank.ac.uk/).

## Acknowledgements

This work was carried out using the computational facilities of the Advanced Computing Research Centre, University of Bristol -http://www.bristol.ac.uk/acrc/.

We are extremely grateful to all the families who took part in this study, the midwives for their help in recruiting them, and the whole ALSPAC team, which includes interviewers, computer and laboratory technicians, clerical workers, research scientists, volunteers, managers, receptionists and nurses.

## Funding

The UK Medical Research Council and Wellcome (Grant ref: 217065/Z/19/Z) and the University of Bristol provide core support for ALSPAC. This publication is the work of the authors and DC will serve as guarantor for the contents of this paper. A comprehensive list of grants funding is available on the ALSPAC website (http://www.bristol.ac.uk/alspac/external/documents/grant-acknowledgements.pdf); This research was specifically funded by Wellcome Trust and MRC (core) (Grant ref: 76467/Z/05/Z), MRC (Grant ref: MR/L022206/1) and Wellcome Trust (Grant ref: 8426812/Z/07/Z).

This work was supported in part by the GW4 BIOMED DTP (D.C., MR/N0137941/), awarded to the Universities of Bath, Bristol, Cardiff and Exeter from the Medical Research Council (MRC)/UKRI.

GMK acknowledges funding support from the Wellcome Trust (grant code: 201486/Z/16/Z), the MQ: Transforming Mental Health (grant code: MQDS17/40), the Medical Research Council UK (grant code: MC_PC_17213 and MR/S037675/1) and the BMA Foundation (J Moulton grant 2019).

AF, GK work in the MRC Integrative Epidemiology Unit (MC_UU_00011)

UK Biobank is generously supported by its founding funders the Wellcome Trust and UK Medical Research Council, as well as the British Heart Foundation, Cancer Research UK, Department of Health, Northwest Regional Development Agency and Scottish Government. The organisation has over 150 dedicated members of staff, based in multiple locations across the UK.

TM is supported by a fellowship from Murdoch Children’s Research Institute.

DB is supported by a National Health and Medical Research (Australia) Investigator Grant (GTN1175744). Research at the Murdoch Children’s Research Institute is supported by the Victorian Government’s Medical Research Operational Infrastructure research Program.

## Author Contribution

Conceptualization: DCPC, AF, SH

Methodology: DCPC, AF, SH, GK

Investigation: DCPC

Supervision: AF, SH, GK

Writing original draft: DCPC

Writing - review and editing: DCPC, AF, SH, GK, DB, TM

## References

1. Hannoodee S, Nasuruddin DN. Acute Inflammatory Response. StatPearls. Treasure Island (FL): StatPearls Publishing Copyright © 2023, StatPearls Publishing LLC.; 2023.

2. Kumar V, Abbas AK, Aster JC. Robbins basic pathology e-book: Elsevier Health Sciences; 2017.

3. Medzhitov RJN. Origin and physiological roles of inflammation. Nature 2008; 454: 428–35.

4. Furman D, Campisi J, Verdin E, et al. Chronic inflammation in the etiology of disease across the life span. Nat Med 2019; 25: 1822–32.

5. Matzinger PJs. The danger model: a renewed sense of self. Science translational medicine 2002; 296: 301–5.

6. Franceschi C, Campisi J. Chronic Inflammation (Inflammaging) and Its Potential Contribution to Age-Associated Diseases. The Journals of Gerontology Series A: Biological Sciences and Medical Sciences 2014; 69: S4–S9.

7. Casimir GJ, Lefèvre N, Corazza F, Duchateau J. Sex and inflammation in respiratory diseases: a clinical viewpoint. Biology of Sex Differences 2013; 4: 16.

8. Gannon CJ, Pasquale M, Tracy JK, McCarter RJ, Napolitano LM. Male gender is associated with increased risk for postinjury pneumonia. Shock 2004; 21: 410–4.

9. Klein SL, Flanagan KL. Sex differences in immune responses. Nat Rev Immunol 2016; 16: 626–38.

10. Deverts DJ, Cohen S, Kalra P, Matthews KA. The prospective association of socioeconomic status with C-reactive protein levels in the CARDIA study. Brain, behavior, and immunity 2012; 26: 1128–35.

11. De Heredia FP, Gómez-Martínez S, Marcos A. Obesity, inflammation and the immune system. Proceedings of the Nutrition Society 2012; 71: 332–8.

12. Straub RH, Hense HW, Andus T, SchoCLmerich J, Riegger GAJ, Schunkert H. Hormone Replacement Therapy and Interrelation between Serum Interleukin-6 and Body Mass Index in Postmenopausal Women: A Population-Based Study. The Journal of Clinical Endocrinology & Metabolism 2000; 85: 1340–4.

13. Fontana L, Eagon JC, Trujillo ME, Scherer PE, Klein S. Visceral Fat Adipokine Secretion Is Associated With Systemic Inflammation in Obese Humans. Diabetes 2007; 56: 1010–3.

14. Wang HJ. Alcohol, inflammation, and gut-liver-brain interactions in tissue damage and disease development. World Journal of Gastroenterology 2010; 16: 1304.

15. Calder PC, Albers R, Antoine J-M, et al. Inflammatory disease processes and interactions with nutrition. British Journal of Nutrition 2009; 101: 1–45.

16. Calabrò P, Golia E, Yeh ET. CRP and the risk of atherosclerotic events. Seminars in immunopathology; 2009: Springer; 2009. p. 79–94.

17. Calabro P, Golia E, TH Yeh EJCpb. Role of C-reactive protein in acute myocardial infarction and stroke: possible therapeutic approaches. Current pharmaceutical biotechnology 2012; 13: 4–16.

18. Sabaawy HE, Ryan BM, Khiabanian H, Pine SR. JAK/STAT of all trades: linking inflammation with cancer development, tumor progression and therapy resistance. Carcinogenesis 2021; 42: 1411–9.

19. Rothwell PM, Fowkes FGR, Belch JF, Ogawa H, Warlow CP, Meade TW. Effect of daily aspirin on long-term risk of death due to cancer: analysis of individual patient data from randomised trials. The Lancet 2011; 377: 31–41.

20. Rothwell PM, Wilson M, Price JF, Belch JF, Meade TW, Mehta Z. Effect of daily aspirin on risk of cancer metastasis: a study of incident cancers during randomised controlled trials. The Lancet 2012; 379: 1591–601.

21. Chu WM. Tumor necrosis factor. Cancer Lett 2013; 328: 222–5.

22. Dowlati Y, Herrmann N, Swardfager W, et al. A meta-analysis of cytokines in major depression. Biol Psychiatry 2010; 67: 446–57.

23. Connelly MA, Gruppen EG, Wolak-Dinsmore J, et al. GlycA, a marker of acute phase glycoproteins, and the risk of incident type 2 diabetes mellitus: PREVEND study. Clin Chim Acta 2016; 452: 10–7.

24. Kinney JW, Bemiller SM, Murtishaw AS, Leisgang AM, Salazar AM, Lamb BT. Inflammation as a central mechanism in Alzheimer’s disease. Alzheimer’s & Dementia: Translational Research & Clinical Interventions 2018; 4: 575–90.

25. Fries GR, Walss-Bass C, Bauer ME, Teixeira AL. Revisiting inflammation in bipolar disorder. Pharmacology Biochemistry and Behavior 2019; 177: 12–9.

26. Oliveira J, OliveiraLMaia A, Tamouza R, Brown A, Leboyer M. Infectious and immunogenetic factors in bipolar disorder. Acta Psychiatrica Scandinavica 2017; 136: 409–23.

27. Kaptoge S, Seshasai SRK, Gao P, et al. Inflammatory cytokines and risk of coronary heart disease: new prospective study and updated meta-analysis. European heart journal 2014; 35: 578–89.

28. Bassuk SS, Rifai N, Ridker PM. High-sensitivity C-reactive protein: clinical importance. Curr Probl Cardiol 2004; 29: 439–93.

29. Gulhar R, Ashraf MA, Jialal I. Physiology, Acute Phase Reactants. StatPearls. Treasure Island (FL): StatPearls Publishing Copyright © 2022, StatPearls Publishing LLC.; 2022.

30. Del Giudice M, Gangestad SWJB, behavior,, immunity. Rethinking IL-6 and CRP: Why they are more than inflammatory biomarkers, and why it matters. Brain, Behavior, and Immunity 2018; 70: 61–75.

31. Scott, Würtz P, Artika, et al. The Biomarker GlycA Is Associated with Chronic Inflammation and Predicts Long-Term Risk of Severe Infection. Cell Systems 2015; 1: 293‱301.

32. Gouin J-P, Glaser R, Malarkey WB, Beversdorf D, Kiecolt-Glaser J. Chronic stress, daily stressors, and circulating inflammatory markers. Health Psychology 2012; 31: 264.

33. Würtz P, Kangas AJ, Soininen P, Lawlor DA, Davey Smith G, Ala-Korpela M. Quantitative Serum Nuclear Magnetic Resonance Metabolomics in Large-Scale Epidemiology: A Primer on -Omic Technologies. American journal of epidemiology 2017; 186: 1084–96.

34. Soininen P, Kangas AJ, Würtz P, Suna T, Ala-Korpela M. Quantitative Serum Nuclear Magnetic Resonance Metabolomics in Cardiovascular Epidemiology and Genetics. Circulation: Cardiovascular Genetics 2015; 8: 192–206.

35. Otvos JD, Shalaurova I, Wolak-Dinsmore J, et al. GlycA: A Composite Nuclear Magnetic Resonance Biomarker of Systemic Inflammation. Clinical Chemistry 2015; 61: 714–23.

36. Chiesa ST, Charakida M, Georgiopoulos G, et al. Glycoprotein Acetyls: A Novel Inflammatory Biomarker of Early Cardiovascular Risk in the Young. Journal of the American Heart Association 2022; 11.

37. Connelly MA, Otvos JD, Shalaurova I, Playford MP, Mehta NN. GlycA, a novel biomarker of systemic inflammation and cardiovascular disease risk. J Transl Med 2017; 15: 219.

38. Kevat AC, Carzino R, Vidmar S, Ranganathan S. Glycoprotein A as a biomarker of pulmonary infection and inflammation in children with cystic fibrosis. Pediatric Pulmonology 2020; 55: 401–6.

39. McGarrah RW, Kelly JP, Craig DM, et al. A novel protein glycan–derived inflammation biomarker independently predicts cardiovascular disease and modifies the association of HDL subclasses with mortality. Clinical Chemistry 2017; 63: 288–96.

40. Dierckx T, Chiche L, Daniel L, Lauwerys B, Van Weyenbergh J, Jourde-Chiche N. Serum GlycA level is elevated in active systemic lupus erythematosus and correlates to disease activity and lupus nephritis severity. Journal of clinical medicine 2020; 9: 970.

41. Akinkuolie AO, Buring JE, Ridker PM, Mora S. A novel protein glycan biomarker and future cardiovascular disease events. Journal of the American Heart Association 2014; 3: e001221.

42. Boyd A, Golding J, Macleod J, et al. Cohort Profile: the ‘children of the 90s’--the index offspring of the Avon Longitudinal Study of Parents and Children. Int J Epidemiol 2013; 42: 111–27.

43. Fraser A, Macdonald-Wallis C, Tilling K, et al. Cohort Profile: the Avon Longitudinal Study of Parents and Children: ALSPAC mothers cohort. Int J Epidemiol 2013; 42: 97–110.

44. Northstone K, Lewcock M, Groom A, et al. The Avon Longitudinal Study of Parents and Children (ALSPAC): an update on the enrolled sample of index children in 2019. Wellcome Open Res 2019; 4: 51.

45. Lewis G, Pelosi AJ, Araya R, Dunn G. Measuring psychiatric disorder in the community: a standardized assessment for use by lay interviewers. Psychol Med 1992; 22: 465–86.

46. Ritchie S, Würtz P, Artika, et al. The Biomarker GlycA Is Associated with Chronic Inflammation and Predicts Long-Term Risk of Severe Infection. Cell Systems 2015; 1: 293–301.

47. Connelly MA, Otvos JD, Shalaurova I, Playford MP, Mehta NN. GlycA, a novel biomarker of systemic inflammation and cardiovascular disease risk. Journal of Translational Medicine 2017; 15.

48. Levine JA, Han JM, Wolska A, et al. Associations of GlycA and high-sensitivity C-reactive protein with measures of lipolysis in adults with obesity. J Clin Lipidol 2020; 14: 667–74.

