## Supplementary file for "Comparison of the stability of Glycoprotein Acetyls and high sensitivity C-reactive protein as markers of chronic inflammation"

| **Contents Page** | |
| --- | --- |
| **Avon Longitudinal Study of Parents and Offspring (ALSPAC)** | **2** |
| **Sample Processing (ALSPAC)** | **4** |
| **Biomarkers of inflammation (ALSPAC)** | **5** |
| **Potential determinants of inflammation (ALSPAC)** | **6** |
| **UK Biobank (UKB)** | **7** |
| **Sample processing (UKB)** | **8** |
| **Potential determinants of inflammation (UKB)** | **9** |
| **Statistical analysis** | **10** |
| **Correlations excluding individuals whom reported an infection** | **11** |
| **Associations with potential determinants of inflammation** | **12** |
| **Supplementary tables** | **13** |
| **Supplementary figures** |  |

**ALSPAC**

Pregnant women resident in Avon, UK with expected dates of delivery between 1st April 1991 and 31st December 1992 were invited to take part in the study. 20,248 pregnancies have been identified as being eligible and the initial number of pregnancies enrolled was 14,541. Of the initial pregnancies, there was a total of 14,676 foetuses, resulting in 14,062 live births and 13,988 children who were alive at 1 year of age. When the oldest children were approximately 7 years of age, an attempt was made to bolster the initial sample with eligible cases who had failed to join the study originally. As a result, when considering variables collected from the age of seven onwards (and potentially abstracted from obstetric notes) there are data available for more than the 14,541 pregnancies mentioned above: The number of new pregnancies not in the initial sample (known as Phase I enrolment) that are currently represented in the released data and reflecting enrolment status at the age of 24 is 906, resulting in an additional 913 children being enrolled (456, 262 and 195 recruited during Phases II, III and IV respectively). The phases of enrolment are described in more detail in the cohort profile paper and its update (see footnote 5 below). The total sample size for analyses using any data collected after the age of seven is therefore 15,447 pregnancies, resulting in 15,658 foetuses. Of these 14,901 children were alive at 1 year of age. Of the original 14,541 initial pregnancies, 338 were from a woman who had already enrolled with a previous pregnancy, meaning 14,203 unique mothers were initially enrolled in the study. As a result of the additional phases of recruitment, a further 630 women who did not enrol originally have provided data since their child was 7 years of age. This provides a total of 14,833 unique women (G0 mothers) enrolled in ALSPAC as of September 2021. The ALSPAC website contains details of all the data that is available through a fully searchable data dictionary and variable search tool http://www.bristol.ac.uk/alspac/researchers/our-data/. Informed consent for the use of data collected via questionnaires and clinics was obtained from participants following the recommendations of the ALSPAC Ethics and Law Committee at the time. *Ethical approval for the collection of biological samples was obtained from the obtained via the ALSPAC Ethics and Law Committee and Local Research Ethics Committees*. Consent for biological samples was obtained in accordance with the Human Tissue Act (2004). Study data were collected and managed using REDCap electronic data capture tools hosted at the University of Bristol. REDCap (Research Electronic Data Capture) is a secure, web-based software platform designed to support data capture for research studies ^1^.

**Sample processing**

Participants fasted overnight (or >6 hours if being seen in the afternoon) before attending the clinic for venepuncture. Blood samples were immediately spun, frozen and stored at -80°C and slowly thawed in a refrigerator (+4°C) prior to analysis processing. The samples were mixed and spun in a centrifuge at 3400 x *g* to remove precipitation. There were no other freeze-thaw cycles and samples were analysed within 3–9 months of collection. Plasma GlycA was quantified as part of a high-throughput proton (1H) NMR metabolomic trait platform ^2^ (Nightingale, UK). Serum hsCRP was measured by automated particle-enhanced immunoturbidimetric assay (Roche UK, Welwyn Garden City, UK). Full details regarding sample processing, NMR analysis, and data processing have been provided elsewhere [38-40]. ALSPAC removed hsCRP values <0.15 mg/L as they were below the detection limit (0.15 mg/L) and two hsCRP measures > 80 mg/L were removed as deemed anomalies. Measures of GlycA ranged from 0.84 to 2.25 mmol/L and measures of serum hsCRP ranged from 0.1 to 80 mg/L. HsCRP was converted from mg/L to mmol/L so that results were presented in the same units as GlycA.

**Biomarkers of inflammation**

In the maternal cohort, we created combined variables separately for the GlycA and hsCRP measures. When present, the measure of GlycA or hsCRP from the 47y clinic was prioritised, but if missing, the measure from the 50y clinic was used (4.69% of participants for GlycA, 3.78% for hsCRP). In this way, we were able to increase sample size (GlycA n=4453 and hsCRP n=4615) and statistical power. The corresponding mean age for the combined GlycA variable was 47.57 years (SD: 4.52) and for the combined hsCRP variable was 47.54 years (SD: 4.51).

**Potential determinants of inflammation**

Mothers’ own and offspring childhood SEP were indexed using maternal self-reported highest education qualification at 8-42 weeks gestation (grouped into college or university degree; A-levels; O-Levels; or NVQs, CSEs or vocational qualification; and household social class, created from the highest social occupation of either parent (dichotomised into manual vs non-manual occupation). Maternal and offspring age (in months) and BMI were recorded at each clinic (BMI indexed as weight in kilogrammes divided by height in meters squared, where weight was measured with the use of Tanita scales to the nearest 0.1 kg and height was measured using a Haroenden standiometer to the nearest 0.1 cm). Offspring sex was recorded and smoking frequency and drinking frequency were self-reported (each categorised as never, infrequently (< weekly) or frequently (≥weekly). Symptoms of infection (including chest infection, cold or fever) in the three weeks prior to attending the clinic (yes/no) were recorded at mean ages 18y and 24y. A doctor’s diagnosis of asthma or eczema was noted at ages 7y and 15y and used to create an atopy diagnosis variable with individuals classified as “none”, “asthma or eczema present” and “both asthma and eczema present”. If there was a discrepancy in answers between the two ages, the affirmative answer was used. A similar method was used for the mother’s atopy variable using confirmation of a doctor’s diagnosis of asthma or eczema at ages 47y and 50y.

**UK Biobank**

Participants were aged 40-70y at baseline, registered with a general practitioner and lived close to 22 assessment centres in England, Scotland, and Wales. Baseline assessments included demographics, lifestyle, and disease history, with linkages to electronic medical records. UK Biobank ethical approval was from the Northwest Multi-centre Research Ethics Committee.

**Sample processing**

Serum hsCRP was measured by automated particle-enhanced immunoturbidimetric assay (Beckman Coulter (UK), Ltd) with detection limits of 0.08 - 80mg/L. Full details regarding hsCRP sample and data processing can be found elsewhere ^3^ and on the UKB website (<https://www.ukbiobank.ac.uk/enable-your-research/about-our-data/biomarker-data>).

**Potential determinants of inflammation**

Self-reported highest education qualification (grouped into college or university degree; A-levels; O-Levels; NVQs, CSEs or vocational qualification; or none of the above) was used as a proxy for SEP. Participants’ sex and age (rounded to whole years) were recorded at baseline where age was a derived variable based on the date of birth and date of attending the initial assessment. BMI was calculated from height and weight measured during the baseline visit (BMI indexed as in ALSPAC and weight was measured with the use of Tanita scales to the nearest 0.1 kg and height was measured using SECA 240 height measure to the nearest 0.1 cm). Participants’ ethnicity was recorded, and we dichotomised categories into white and non-white. Smoking frequency and alcohol intake were self-reported at baseline as never, infrequently (< weekly) or frequently (≥weekly).

The diseases included were those for which participants had received a diagnosis for recorded across inpatient records in either the primary or secondary position from 2006 onwards. Each overarching disease contained multiple types of disease which can be found in table 2 of the supplement along with their ICD-10 code and the number of individuals with the disease. Specifically, we used diagnoses for types of arthritis, types of Crohn’s Disease, types of SLE, MS, types of chronic sinusitis, the different presentations of T2D, hepatitis C, types of eczema, and types of asthma.

**Statistical Analysis**

For analysis using ALSPAC, model 3 included the variables: sex, age, SEP, household social class, atopy, smoking status and drinking frequency. Ethnicity was not adjusted for as non-white participants had been removed. For UKB, when replicating the analysis in ALSPAC model 3 included the variables: age, sex, ethnicity, BMI, SEP, smoking status and drinking frequency. The variables included as covariates when investigating the relationship between ICD-10 diagnosed pro-inflammatory diseases and levels of CRP and GlycA were SEP, sex, BMI, ethnicity and age.

**Correlations excluding individuals who report an infection at any timepoint in the ALSPAC offspring**

There were moderate-to-strong correlations between measures of GlycA taken 5-6 weeks apart at ages 15y (r=0.75; 95% CI=0.56, 0.94), 18y (r=0.74; 0.64, 0.85) and 24y (*r*=0.74; 0.51, 0.98). hsCRP had a strong short-term correlation (*r*=0.77; 0.59, 0.95) at the one-time point for which QC data was available (age 24y).

GlycA had moderate correlations between measures taken years apart in adolescence/early-adulthood (between 15y and 18y: r=0.52; 0.47, 0.56; between 15y and 24y: *r* =0.37; 0.31, 0.44 and; between 18y and 24y: r = 0.43; 0.37, 0.49). These were larger than equivalent correlations of hsCRP (between 15y and 18y: r=0.40; 0.35, 0.46; between 15y and 24y: *r* =0.25; 0.18, 0.32 and; between 18y and 24y: r = 0.32; 0.25, 0.38).

Concurrently measured GlycA and hsCRP levels were moderately correlated at all ages: 15y (r=0.44; 0.40, 0.48), 18y (r=0.55; 0.51, 0.59), 24y (r=0.54; 0.50, 0.59).

**Associations with potential determinants of inflammation**

ALSPAC offspring: Multiple inflammation-related factors at ages 15y, 18y and 24y were associated with both GlycA and hsCRP in a similar pattern. For example, in the unadjusted associations (model 1), higher BMI and lower socioeconomic position (SEP) (mother having a lower level of educational qualification) were positively associated with hsCRP and GlycA at all three time points. Having a recent infection within the three weeks preceding the clinic was positively associated with hsCRP and GlycA at 15y and 18y (the two time points at which recent infection was recorded), and smoking status was positively associated with hsCRP and GlycA at ages 18y and 24y (the two time points it was recorded). The only factor where the two biomarkers diverged was sex. GlycA was higher in females than males at ages 15y and 18y, but this difference attenuated to the null by 24y. Conversely, mean hsCRP did not differ between sexes at 15y but was higher in females than males at both 18y and 24y. In models adjusted for age and sex (model 2), results were similar to those described for model 1.

UKB: In the unadjusted model (model 1) and in the model adjusted for age and sex (model 2) arthritis, asthma, MS, chronic sinusitis, diabetes, Crohn’s disease, and eczema were each associated with both higher GlycA and hsCRP

| **Supplementary Table 1: A breakdown of the variants within each disease, the number of individuals with a diagnosis and what cluster they fall under** | | | | |
| --- | --- | --- | --- | --- |
| Disease | Diagnosis? | N | Variants | Cluster |
| Asthma | Yes | 8410 | - allergic asthma - nonallergic asthma - mixed asthma - unspecified asthma | Atopy |
|  | No | 104014 |  |  |
| Eczema | Yes | 587 | - Other atopic dermatitis - Unspecified atopic dermatitis - Nummular dermatitis - Dyshidrosis - Cutaneous autosensitisation - Infective dermatitis - Erythema intertrigo - Pityriasis alba - Other specified dermatitis - Unspecified dermatitis | Atopy |
|  | No | 111837 |  |  |
| Hepatitis C | Yes | 102 | - Acute hepatitis C - Chronic Viral hepatitis | Chronic Infection |
|  | No | 112322 |  |  |
| Chronic Sinusitis | Yes | 733 | - Chronic Sinusitis | Chronic Infection |
|  | No | 11691 |  |  |
| Arthritis | Yes | 1520 | - Seropositive rheumatoid arthritis - Other rheumatoid arthritis - Juvenile arthritis. | Autoimmune Disease |
|  | No | 110904 |  |  |
| Diabetes | Yes | 883 | Diabetes with:   - Coma - Ketoacidosis - Renal complications - Ophthalmic complications - Neurological complications - Peripheral circulatory complications - Other specified complications - Multiple complications - Unspecified complications - Without complications | Autoimmune Disease |
|  | No | 11541 |  |  |
| Lupus | Yes | 45 | - Discoid lupus erythematosus - Subcutaneous lupus erythematosus - Other local lupus erythematosus | Autoimmune Disease |
|  | No | 112379 |  |  |
| Multiple Sclerosis | Yes | 385 | - Multiple Sclerosis | Autoimmune Disease |
|  | No | 112039 |  |  |
| Crohn’s Disease | Yes | 547 | - Crohn’s disease of the small intestine - Crohn’s disease of the large intestine - Other Crohn’s disease - Unspecified Crohn’s disease | Autoimmune Disease |
|  | No | 111877 |  |  |

| **Supplementary Table 2: Comparison of demographic characteristics in the ALSPAC offspring and mothers** | | | |
| --- | --- | --- | --- |
| Variable | Category | N | Mean (SD) or % for categorical |
| **15y** |  |  |  |
| GlycA (mmol/L) |  | 3294 | 1.21 (0.13) |
| CRP (mg/L) |  | 3374 | 1.22 (3.68) |
| BMI at 15y |  | 3328 | 21.53 (3.54) |
| Age at 15y |  | 3368 | 185.55 (4.11) |
| Sex | Male | 1440 | 48.90 |
|  | Female | 1505 | 51.10 |
| Household Social Class | Non-manual | 2566 | 86.37 |
|  | Manual | 405 | 13.63 |
| Mother highest educational attainment | Degree | 578 | 18.99 |
|  | A levels | 911 | 29.93 |
|  | O levels | 1077 | 35.38 |
|  | NVQs, CSEs, vocational | 478 | 15.70 |
| Infection at 15y | No infection | 2550 | 78.24 |
|  | Infection | 709 | 21.76 |
| Atopy | None | 1369 | 67.64 |
|  | Either | 542 | 26.78 |
|  | Both | 113 | 5.58 |
| **18y** |  |  |  |
| GlycA (mmol/L) |  | 3108 | 1.22 (0.14) |
| CRP (mg/L) |  | 3189 | 1.62 (5.02) |
| BMI at 18y |  | 3109 | 22.73 (3.92) |
| Age at 18y |  | 3189 | 213.39 (4.80) |
| Sex | Male | 1318 | 48.91 |
|  | Female | 1377 | 51.09 |
| Mother highest educational attainment | Degree | 589 | 20.55 |
|  | A levels | 874 | 30.50 |
|  | O levels | 973 | 33.95 |
|  | NVQs, CSEs, vocational | 430 | 15.00 |
| Household Social Class | Non-manual | 2445 | 87.54 |
|  | Manual | 348 | 12.46 |
| Infection at 18y | No infection | 2003 | 78.00 |
|  | Infection | 565 | 22.00 |
| Type of smoker | Non smoker | 1300 | 63.88 |
|  | Infrequent smoker | 290 | 14.25 |
|  | Frequent smoker | 445 | 21.87 |
| Alcohol intake at 18y | Non drinker | 528 | 17.50 |
|  | Infrequent drinker | 1268 | 42.03 |
|  | Frequent drinker | 1221 | 40.47 |
| Presence of atopic disease | None | 1202 | 67.99 |
|  | Either | 460 | 26.02 |
|  | Both | 106 | 6.00 |
| **24y** |  |  |  |
| GlycA (mmol/L) |  | 3108 | 1.22 (0.14) |
| CRP (mg/L) |  |  |  |
| BMI at 18y |  | 2372 | 21.28 (3.42) |
| Age at 18y |  | 2455 | 213.21 (4.86) |
| Sex | Male | 1063 | 41.46 |
|  | Female | 1501 | 58.54 |
| Mother highest educational attainment | Degree | 583 | 20.98 |
|  | A levels | 849 | 30.55 |
|  | O levels | 960 | 34.54 |
|  | NVQs, CSEs, vocational | 387 | 13.93 |
| Household Social Class | Non-manual | 2418 | 89.16 |
|  | Manual | 294 | 10.84 |
| Presence of atopic disease | None | 1100 | 66.03 |
|  | Either | 466 | 27.97 |
|  | Both | 100 | 6.00 |
| Type of smoker at 24y | Non smoker | 1116 | 36.19 |
|  | Infrequent smoker | 1417 | 45.95 |
|  | Frequent smoker | 551 | 17.87 |
| Alcohol intake at 24y | Non drinker | 43 | 1.83 |
|  | Infrequent drinker | 1644 | 70.08 |
|  | Regular drinker | 659 | 28.09 |
| **47y** |  |  |  |
| GlycA (mmol/L) |  | 4243 | 1.24 (0.16) |
| CRP (mg/L) |  | 4446 | 2.22 (3.76) |
| BMI at 47y |  | 4292 | 26.54 (5.28) |
| Age at 47y |  | 4305 | 47.41 (4.47) |
| Highest educational attainment | Degree | 781 | 19.61 |
|  | A levels | 1199 | 30.11 |
|  | O levels | 1399 | 35.13 |
|  | NVQs, CSEs, vocational | 603 | 15.14 |
| Grandparents household social class | Non-manual | 2410 | 70.82 |
|  | Manual | 993 | 29.18 |
| **50y** |  |  |  |
| GlycA (mmol/L) |  | 2709 | 1.25 (0.17) |
| CRP (mg/L) |  | 2584 | 2.23 (4.13) |
| BMI at 47y |  | 2721 | 26.27 (5.15) |
| Age at 47y |  | 2730 | 50.32 (4.41) |
| Highest educational attainment | Degree | 559 | 22.49 |
|  | A levels | 793 | 31.90 |
|  | O levels | 861 | 34.63 |
|  | NVQs, CSEs, vocational | 273 | 10.98 |
| Grandparents household social class | Non-manual | 1619 | 73.03 |
|  | Manual | 598 | 26.97 |

| **Supplementary Table 3: Comparison of demographic characteristics in UKB** | | | |
| --- | --- | --- | --- |
| Variable | Category | N | Mean (SD) or % for categorical |
| GlycA (mmol/L) |  | 112424 | 0.79 (0.12) |
| CRP (mg/L) |  | 112424 | 2.61 (4.42) |
| Ethnicity | White | 98984 | 88.59 |
|  | Non-white | 12745 | 11.41 |
| BMI at 18y |  | 12424 | 22.73 (4.77) |
| Age (years) |  | 112424 | 56.51 (8.09) |
| Sex | Male | 51530 | 45.84 |
|  | Female | 60894 | 54.16 |
| Mother highest educational attainment | Degree | 36289 | 32.67 |
|  | A levels | 12329 | 11.10 |
|  | O levels | 23726 | 21.26 |
|  | NVQs, CSEs, vocational | 19539 | 17.59 |
|  | None of the above | 19186 | 17.27 |
| Diagnosis of disease | No diagnosis | 95639 | 45.97 |
|  | Diagnosis | 112424 | 54.03 |
| Type of smoker | Non smoker | 61060 | 54.59 |
|  | Infrequent smoker | 38929 | 34.80 |
|  | Frequent smoker | 11864 | 10.60 |
| Alcohol intake | Non drinker | 4902 | 4.37 |
|  | Infrequent drinker | 4055 | 3.62 |
|  | Frequent drinker | 103195 | 92.01 |

| **Supplementary Table 4: Cross-sectional associations between key determinants of inflammation and GlycA in UKB excluding individuals with pro-inflammatory diseases** | | | | | | | | | |
| --- | --- | --- | --- | --- | --- | --- | --- | --- | --- |
| **Variable** | **Model 1** | | | **Model 2 *** | | | **Model 3**** | **(N=23973)** |  |
|  | Beta (se) | 95% CI | *p* | Beta (se) | 95% CI | *p* | Beta (se) | 95% CI | *p* |
| **GlycA** |  |  |  |  |  |  |  |  |  |
| BMI at (kg/m^2^) | 1.55 (0.01) | 1.52, 1.57 | <0.001 | 1.55 (0.01) | 0.52, 1.58 | <0.001 | 1.60 (0.03) | 1.55, 1.66 | <0.001 |
| Age (months) | 0.94 (0.03) | 0.89, 0.99 | <0.001 | 0.94 (0.03) | 0.90, 1.00 | <0.001 | 0.72 (0.05) | 0.61, 0.83 | <0.001 |
| Sex (female) | 0.0002 (0.002) | -0.003, 0.003 | 0.905 | -0.001 (0.002) | -0.004, 0.003 | 0.728 | -0.01 (0.004) | -0.02, -0.01 | <0.001 |
| Highest educational attainment (None recorded) | 0.23 (0.005) | 0.22, 0.24 | <0.001 | 0.20 (0.004) | 0.19, 0.21 | <0.001 | 0.10 (0.01) | 0.08, 0.11 | <0.001 |
| Smoking frequency | 0.02 (0.002) | 0.02, 0.07 | <0.001 | 0.02 (0.002) | 0.02, 0.03 | <0.001 | 0.03 (0.002) | 0.02, 0.03 | <0.001 |
| Alcohol intake frequency | -0.05 (0.002) | -0.06, -0.05 | <0.001 | -0.05 (0.002) | -0.06, -0.05 | <0.001 | -0.03 (0.004) | -0.04, -0.03 | <0.001 |
| **CRP** |  |  |  |  |  |  |  |  |  |
| BMI (kg/m^2^) | 1.95 (0.01) | 1.92, 1.97 | <0.001 | 1.97 (0.01) | 1.94, 1.99 | <0.001 | 1.98 (0.03) | 1.93, 2.03 | <0.001 |
| Age (months) | 0.98 (0.03) | 0.93, 1.03 | <0.001 | 0.98 (0.03) | 0.93, 1.03 | <0.001 | 0.84 (0.06) | 0.73, 0.95 | <0.001 |
| Sex (female) | -0.01 (0.002) | -0.01, -0.006 | <0.001 | -0.01 (0.002) | -0.01, -0.007 | <0.001 | -0.04 (0.004) | -0.04, -0.03 | <0.001 |
| Highest educational attainment (None recorded) | 0.22 (0.005) | 0.21, 0.23 | <0.001 | 0.19 (0.005) | 0.18, 0.20 | <0.001 | 0.07 (0.01) | 0.05, 0.09 | <0.001 |
| Smoking frequency | 0.02 (0.002) | 0.016, 0.002 | <0.001 | 0.02 (0.002) | 0.02, 0.03 | <0.001 | 0.03 (0.002) | 0.02, 0.03 | <0.001 |
| Alcohol intake frequency | -0.05 (0.002) | -0.05, -0.04 | <0.001 | -0.05 (0.002) | -0.05, -0.04 | <0.001 | -0.02 (0.004) | -0.03, -0.01 | <0.001 |
| * Adjusted for sex and age, estimates for age were adjusted for sex and vice versa.  ** Adjusted sex, age, SEP, smoking frequency and drinking frequency. The variable was not included as a covariate if used as the exposure. | | | | | | | | | |

Supplementary Figure 1: Participant flowchart for ALSPAC offspring

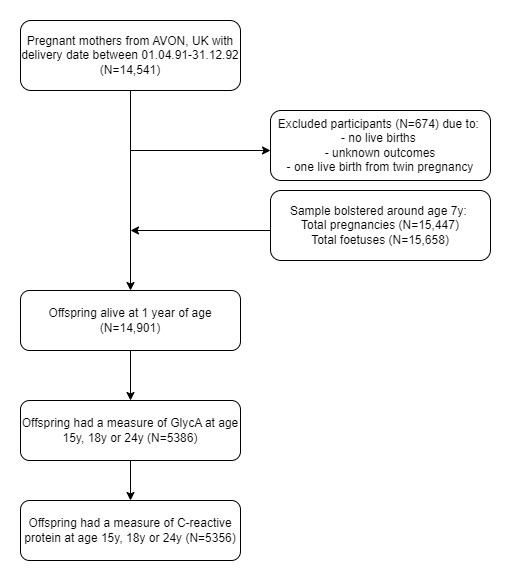

Supplementary Figure 2: Participant flowchart for ALSPAC mothers

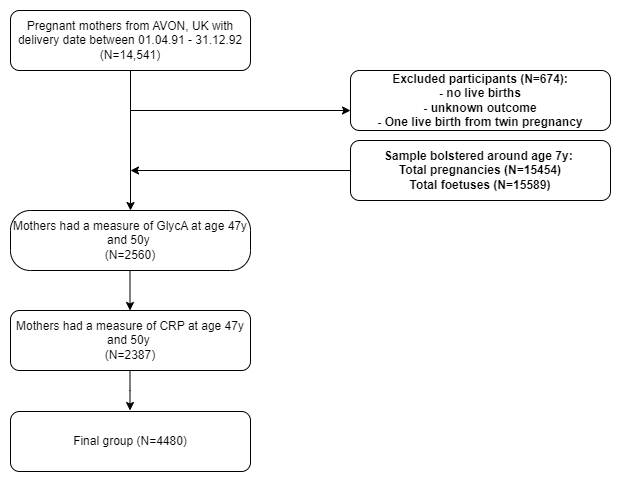

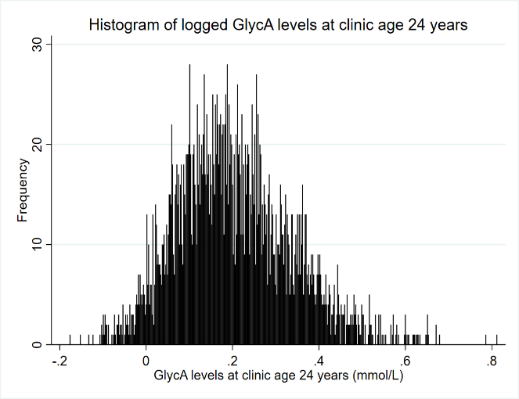

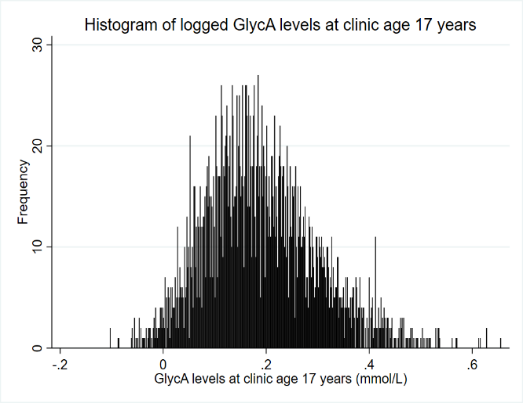

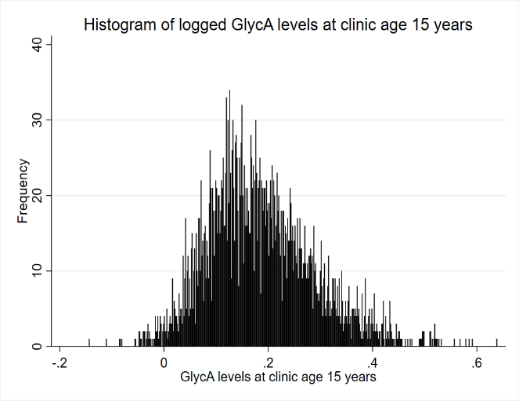

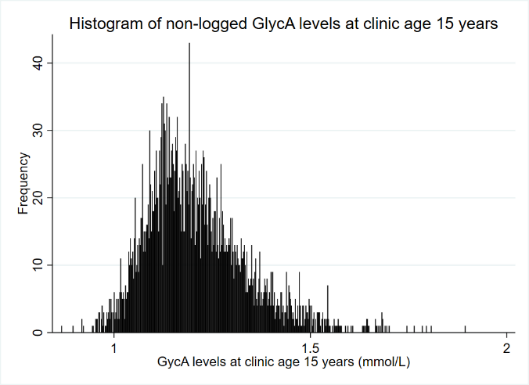

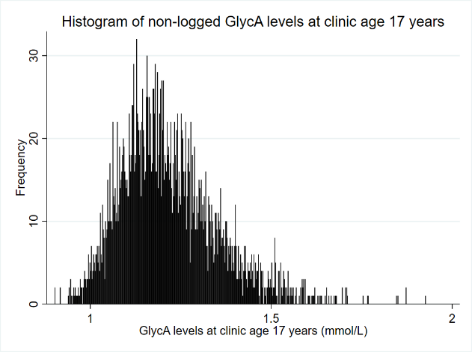

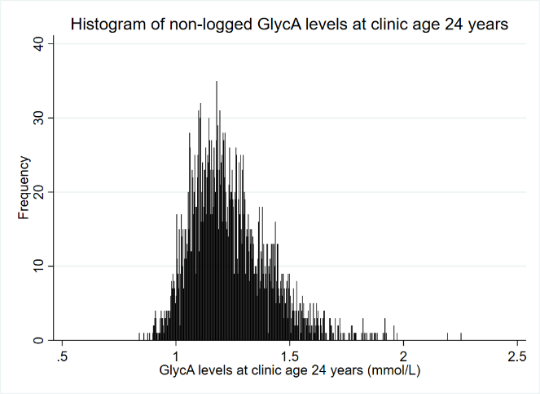

Supplementary Figure 3: distribution of non-logged and logged GlycA measures at 15y, 18y and 24y in ALSPAC offspring and at mean clinic ages 47 years and 50 years

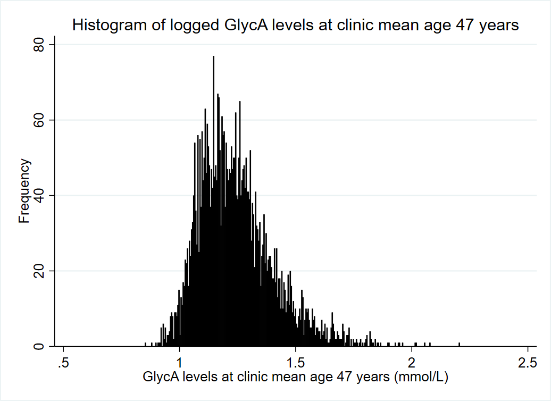

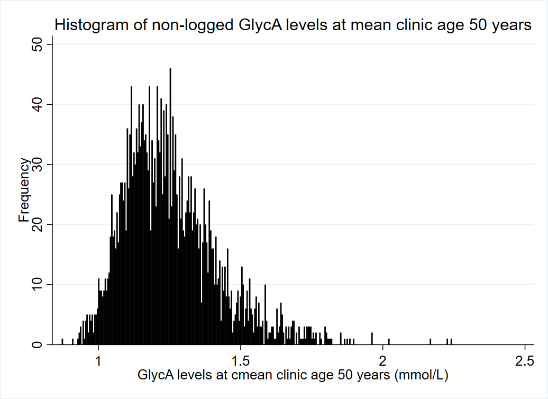

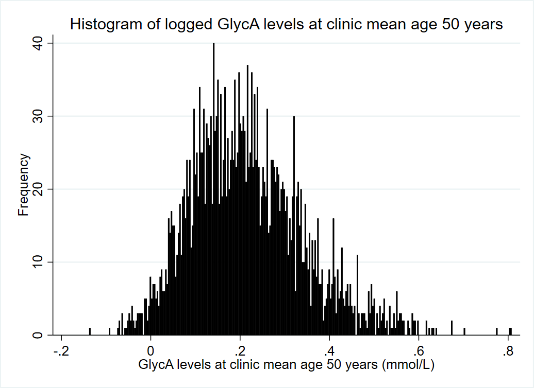

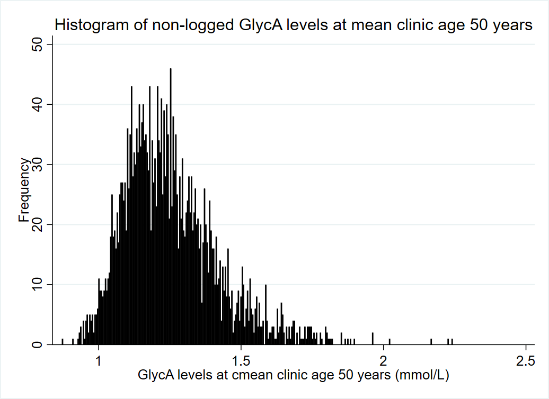

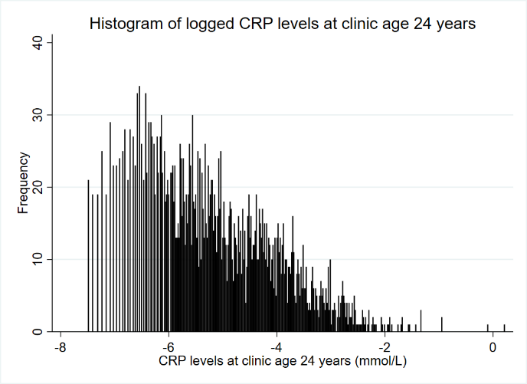

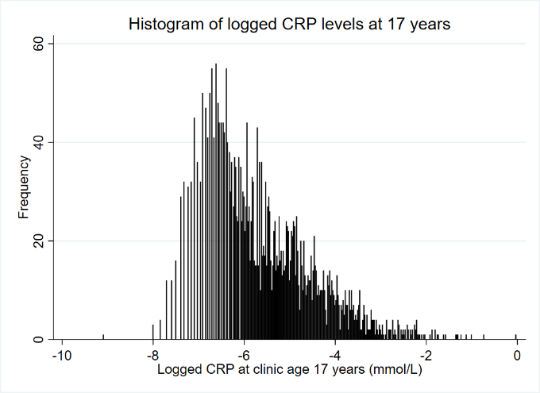

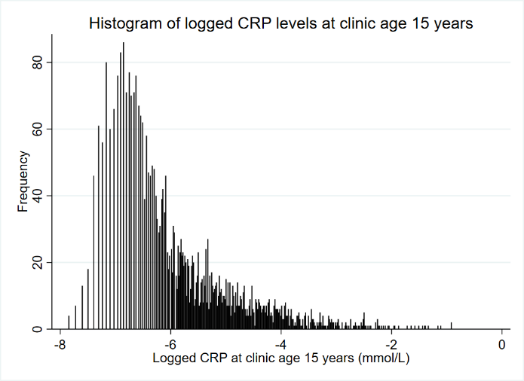

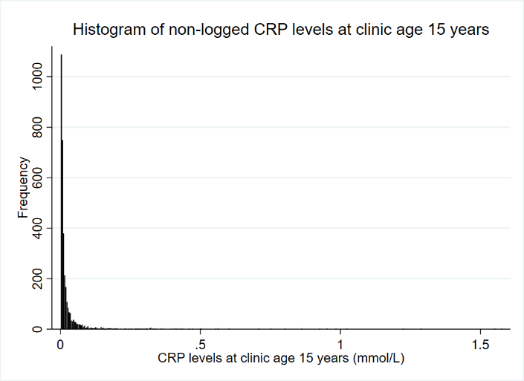

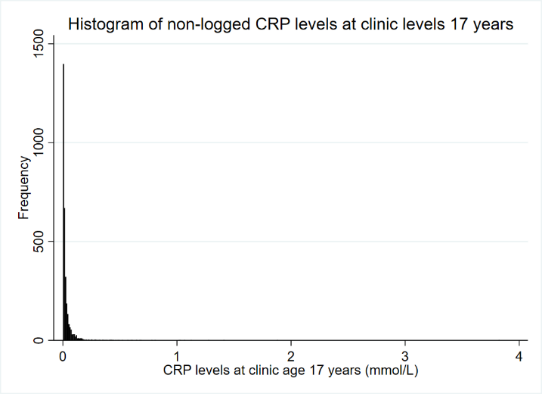

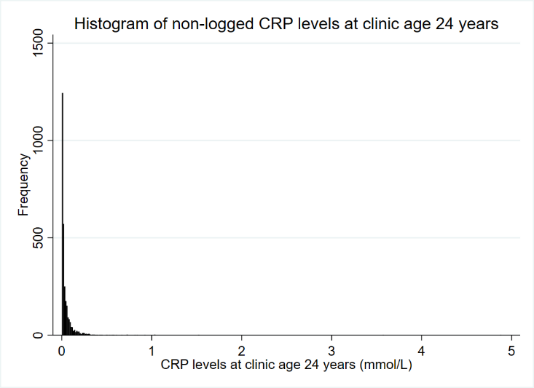

Supplementary Figure 4: Distribution of non-logged and logged hsCRP measures at 15y, 18y and 24y in ALSPAC offspring and mean ages 47 years and 50 years in ALSPAC mothers

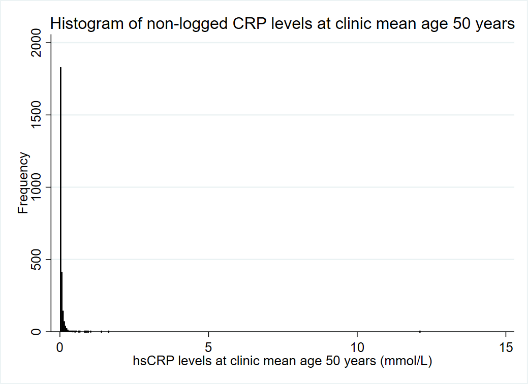

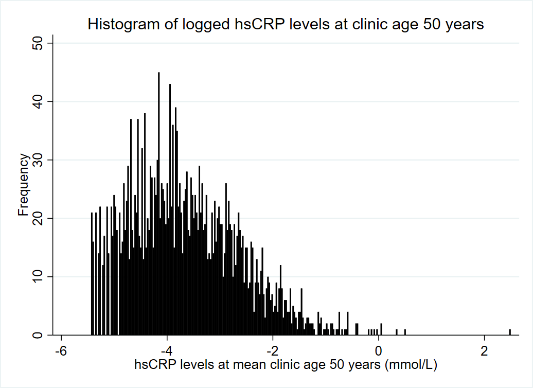

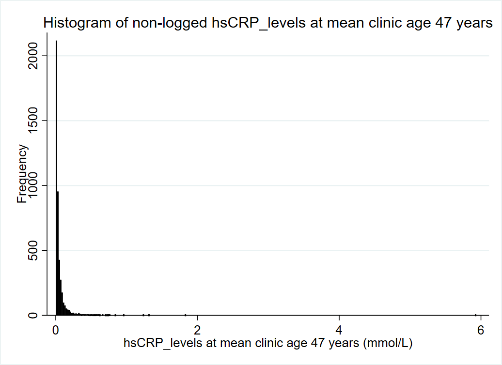

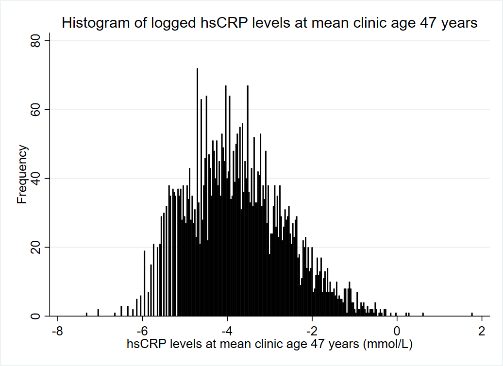

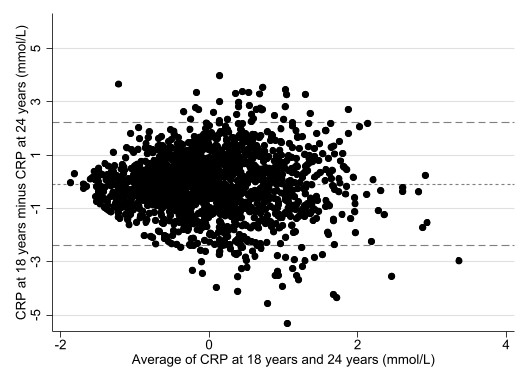

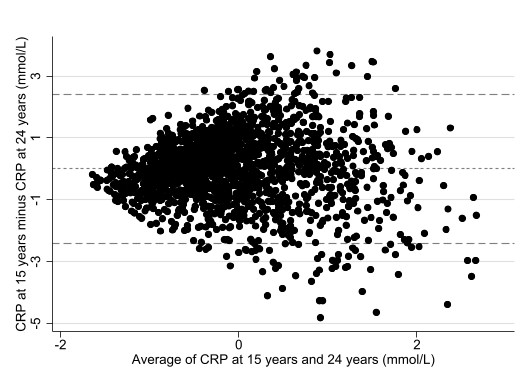

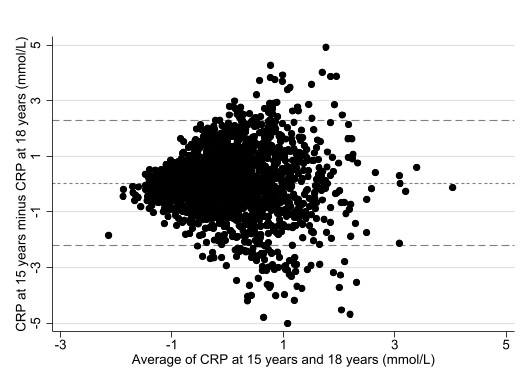

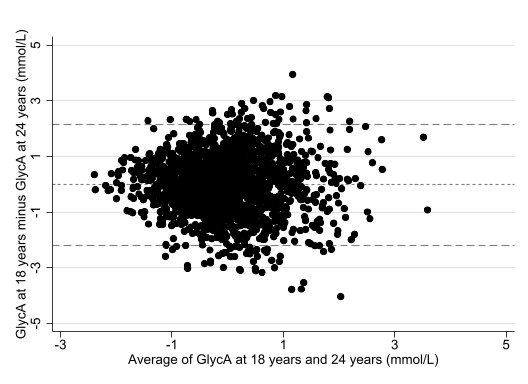

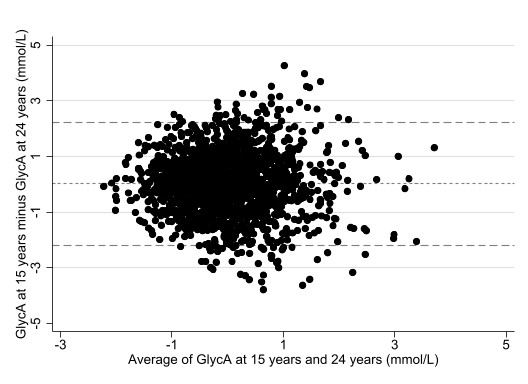

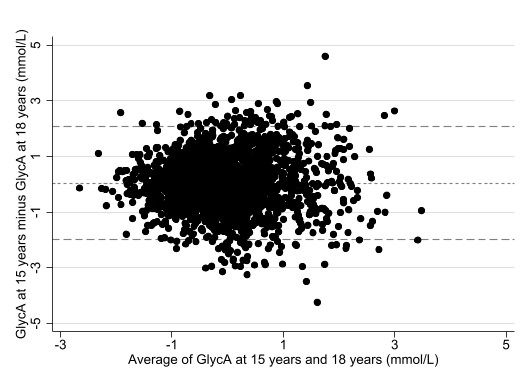

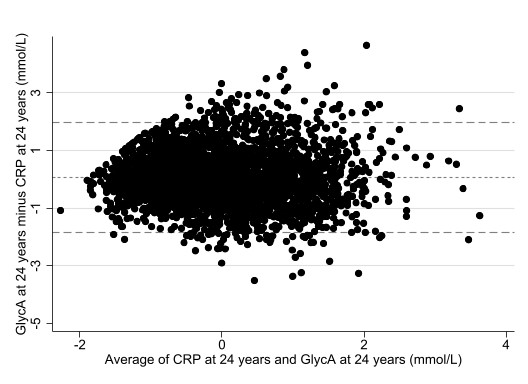

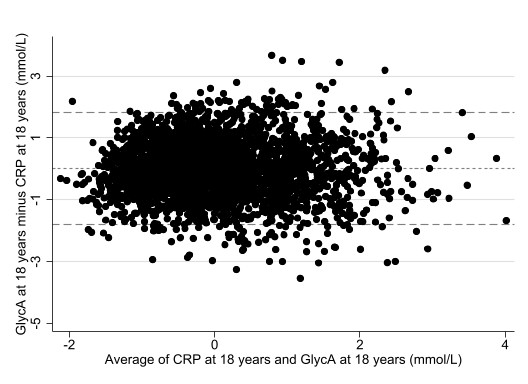

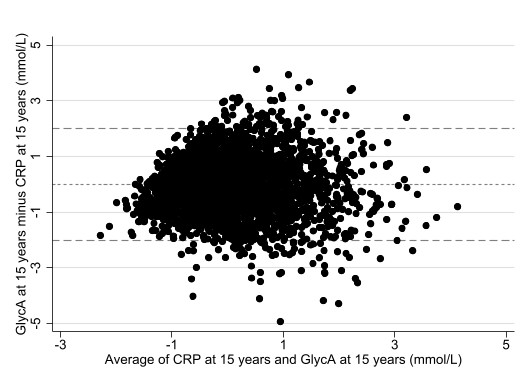

Supplementary Figure 5: Bland-Altman plots of GlycA and CRP measures at different timepoints using ALSPAC offspring data

*NB: Measures are log-transformed and z-scored*

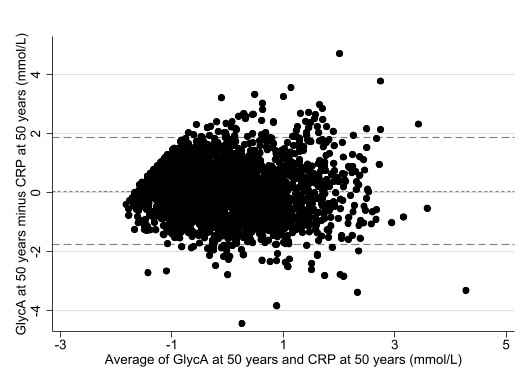

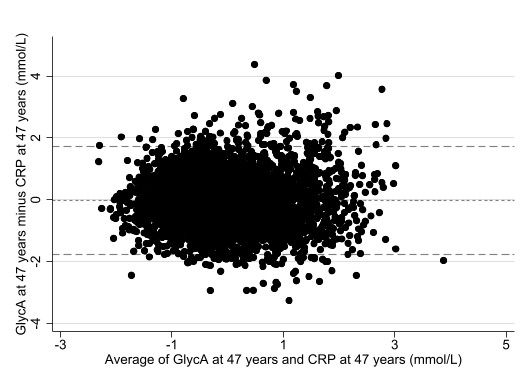

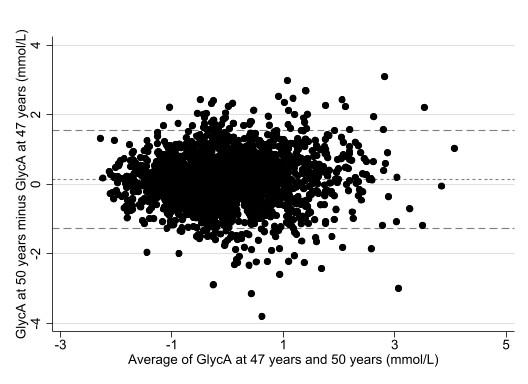

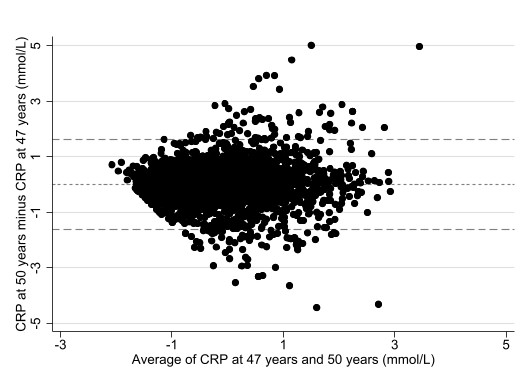

Supplementary Figure 6: Bland-Alman plots of GlycA and CRP measures at different timepoints using ALPSAC mother data

*N.B: Measures are log-transformed and z-scored*
